# Diabetes Advances Cardiomyocyte Senescence through miR-103a-3p/Rnd3/STAT3 Signaling

**DOI:** 10.1101/2024.01.16.24301350

**Authors:** Linxu Wu, Xinglin Zhu, Cai Luo, Yan Chen, Yin Liu, Yangyang Zhao, Shanshan Pan, Kaijia Shi, Jingci Xing, Xuebin Lin, Jianmin Qiu, Shuya Zhang, Jinxuan Chai, Zhihua Shen, Wei Jie, Junli Guo

## Abstract

**BACKGROUND:** Rnd3, a small Rho-GTPase, is involved in various cardiovascular diseases, but its role in diabetes-induced cardiomyocyte senescence is unclear. This study aims to investigate the role of Rnd3 in cardiomyocyte senescence in diabetes cardiomyopathy (DCM).

**METHODS:** *Rnd3* expression was assessed in human tissue using qPCR, and its correlation with clinical parameters was analyzed. *Rnd3* and Senescence-associated secretory phenotype (SASP)-related marker expression was determined in heart tissue from juvenile/mature mice and streptozotocin-induced diabetic Sprague–Dawley rats. H9C2 and AC16 cardiomyocytes were exposed to high glucose ([HG] =35 mmol/L D-glucose), and Rnd3 and SASP-related marker expression was examined. *Rnd3* expression in H9C2 cells was disrupted using CRISPR/Cas9 technology, and cellular senescence of HG-treated H9C2 cells was evaluated. MicroRNA sequencing was conducted on AC16 cells to identify differentially expressed microRNAs regulated by HG. Among these microRNAs, miR-103a-3p was selected for further investigation in a clinical diabetes cohort and to examine its binding to *Rnd3* 3′UTR and its effect on cell senescence. Finally, the role of interaction of Rnd3 and STAT3 in cardiomyocyte senescence was investigated.

**RESULTS:** *Rnd3* mRNA levels were reduced in peripheral blood mononuclear cells in diabetic patients, which was negatively correlated to patients age but positively correlated to cardiac function. Older mice exhibited compromised cardiac function, increased SA-β-gal-positive cells, and SASP marker levels versus younger mice. HG stimulation inhibited Rnd3 expression but enhanced cellular senescence. *Rnd3* knockout resulted in greater cellular senescence on HG-insulted cardiomyocytes. miR-103a-3p was identified as a key HG-regulated microRNA that binds to *Rnd3* 3′UTR. AAV9 carrying miR-103a-3p sponges and *Rnd3*-overexpressing plasmids mitigated cellular senescence in diabetic rats. Interestingly, miR-103a-3p levels positively correlated with blood glucose concentrations. HG stimulation amplified phosphorylated STAT3(Tyr705) in H9C2 cells, which was intensified by *Rnd3* knockout. Rnd3 binds with STAT3 in cytoplasm and promote proteasome-induced ubiquitination of STAT3. S3I-201 mitigated HG-activated STAT3 and cardiomyocyte senescence.

**CONCLUSION:** Diabetes causes cardiomyocyte senescence through miR-103a-3p/Rnd3/STAT3 signaling, suggesting a potential treatment strategy for DCM.

---

The medical and social demographic issues arising from global population aging are becoming increasingly prominent.^1^ According to the World Health Organization, the global population at 65 years and older is projected to rise from 10% in 2022 to 16% in 2050.^2^ This growth is occurring at an unprecedented rate and is expected to accelerate in the coming decades, especially in developing countries. Along with aging, the incidence of diabetes and other chronic diseases is also increasing. In 2021,approximately 537 million adults worldwide had diabetes, and the total number of people with diabetes is expected to increase to 783 million by 2045.^3^ Diabetes alone caused 6.7 million deaths in 2021. Studies have shown a strong correlation between diabetes and aging, particularly in the population older than 65 years, where the prevalence of diabetes is as high as 25%–30%.^4^ Unfortunately, aging and diabetes are considered independent risk factors for cardiovascular disease.^5,6^ Furthermore, diabetes can exacerbate cellular aging, known as pathological senescence. Diabetes exacerbates age-related heart damage through mechanisms, such as fibrosis, NAD^+^ metabolism abnormalities, oxidative stress, and autophagic inhibition.^7–11^ Therefore, reducing cardiac cell senescence exacerbated by diabetes is important in preventing cardiovascular events.

Aging cardiomyocytes exhibit contraction and metabolic dysfunction, and can secrete factors associated with the senescence-associated secretory phenotype (SASP), leading to remodeling of the local cardiac microenvironment.^11–12^ Although various mechanisms can induce cardiomyocyte senescence, metabolic stressors, represented by diabetes, can induce the disease phenotype of cardiomyocytes.^12^ Unlike the senescence of other cells, cardiomyocytes, as terminally differentiated cells, cannot simply be determined as experiencing senescence from cell cycle arrest. The aging of cardiomyocytes shows several features,^13–15^ which was mainly manifested as DNA damage, SASP expression, systolic dysfunction, endoplasmic reticulum stress, cell hypertrophy and mitochondrial dysfunction. Although diabetes has been shown to be an important factor in promoting cardiomyocyte aging, senescent mechanisms and targets in the heart, especially under the diabetic condition, remain unclear.

Rnd3, also known as Rho family GTPase 3, is one of the atypical members of the Rho GTPase superfamily. Rnd3 is an endogenous inhibitor of Rho-associated kinase (ROCK) and plays an important role in cytoskeleton formation, cell proliferation, migration, and differentiation, the cell cycle, and apoptosis.^16,17^ Rnd3 is involved in pathophysiological processes, such as brain edema caused by ependymal cell proliferation, cardiomyocyte apoptosis induced by stress overload, chronic heart failure, post-heart failure angiogenesis, arrhythmia, and post-infarction inflammation.^18–22^ A recent report suggested that cardiac fibroblast-specific Rnd3 activation plays an important protective role in the pathogenesis of diabetic cardiomyopathy (DCM).^23^ These reports highlight the potential targeted value of Rnd3 in the treatment of cardiovascular disease.

This study mainly aimed to investigate the mechanistic role of Rnd3 in cardiomyocyte senescence under the diabetic condition. Our results show a novel role of dysregulation of Rnd3 in diabetes-induced cardiomyocyte senescence and suggest an alternative treatment strategy for DCM.

## METHODS

For expanded methods, please see the Supplemental Material.

## RESULTS

### *Rnd3* Expression in Human Tissue

To clarify the relationship between *Rnd3* and diabetes, we first searched the Gene Expression Omnibus (GEO) database for T2DM data. We found that *Rnd3* mRNA expression levels in peripheral blood mononuclear cells (GSE23561, GSE95849) and coronary tissue (GSE13760) in the T2DM population were significantly lower than those in the non-diabetic control groups (Fig. 1a). Then, used our cohort, we found that *Rnd3* mRNA expression in peripheral blood mononuclear cells of T2DM patients was significantly lower compared with that in controls (Fig. 1b, Table S1). Correlation analysis showed that *Rnd3* mRNA expression was significantly negatively correlated with age (r=−0.546, *P*=0.001), FBG (fasting blood glucose, r=-0.368, *P*=0.0497), cardiac troponin I (cTnI) (r=−0.4666, *P*=0.0062), myoglobin (r=−0.6429, *P*<0.0001), NT-proBNP(r=−0.4087, *P*=0.0224), but positively correlated with EF% (r=0.5628, *P*=0.0007) and E/A ratio (r=0.4305, *P*=0.0357) (Fig. 1c). These results suggest that T2DM has an inhibitory effect on *Rnd3*, and that *Rnd3* mRNA expression is related to aging and myocardial injury.

**Figure 1.**
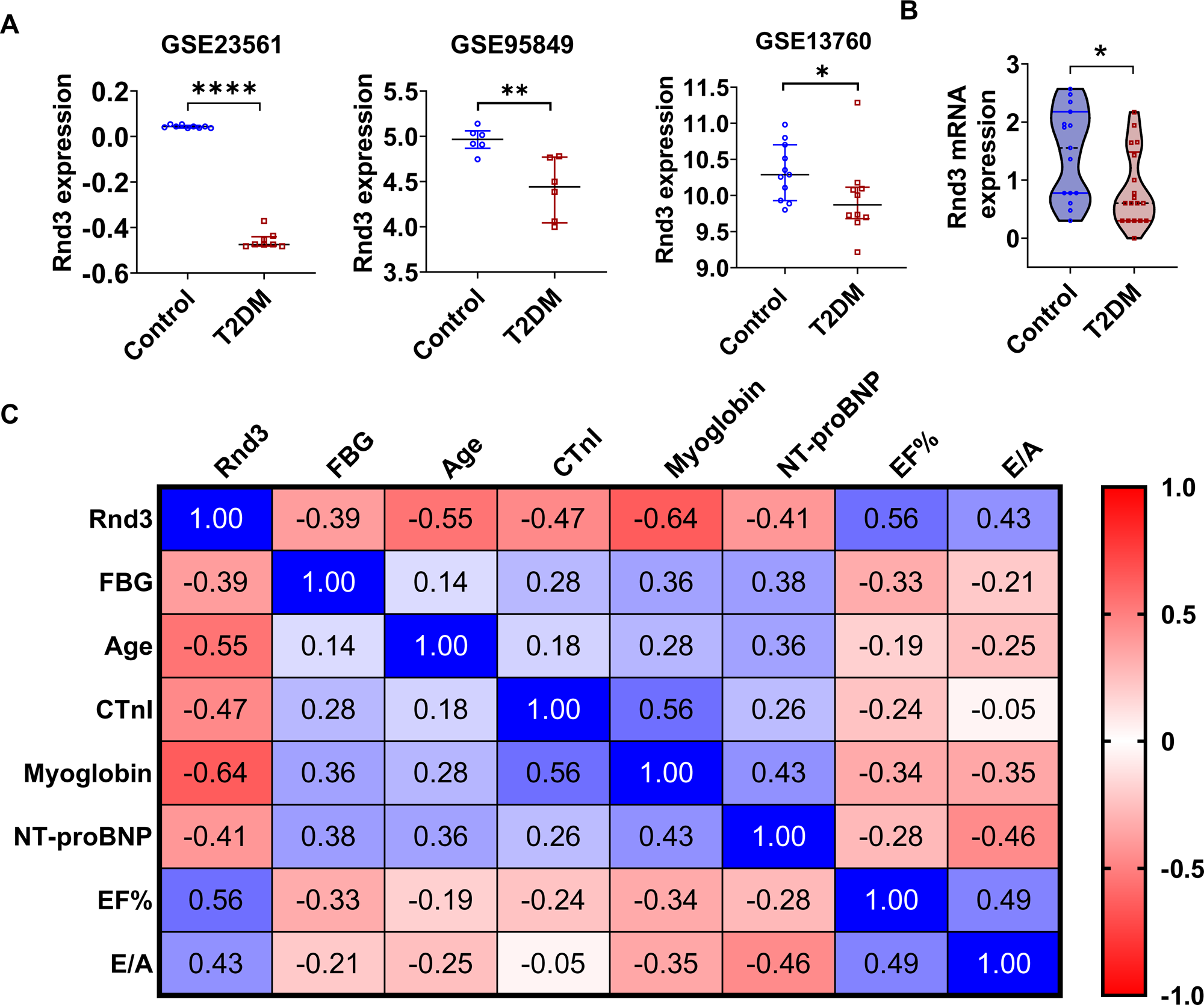
*Rnd3* expression in human tissue based on public databases and a clinical cohort. (**A**) Datasets in Gene Expression Omnibus (GEO) showed that *Rnd3* mRNA levels were enhanced in peripheral blood cells (GSE23561, GSE95849) and coronary artery media (GSE13760) in patients with diabetes compared with controls. Data were analyzed by the Mann–Whitney test; \**P*<0.05, \*\**P*<0.01, \*\*\*\**P*<0.0001. (**B**) *Rnd3* mRNA levels in peripheral blood mononuclear cells of a clinical cohort Diabetic patients and non-diabetic patients. Data were analyzed by the Mann–Whitney test; \**P*<0.05. (**C**) Spearman correlation analysis of *Rnd3* mRNA levels with fasting blood glucose (FBG), age, cardiac troponin I (cTnI), myoglobin, NT-proBNP, ejection fraction (EF%) and E/A ratio in a clinical cohort.

### *Rnd3* Expression and Cardiac Function in Aged Mice

By comparing gene expression in heart tissue and cardiac function in juvenile and aged C56BL6/J mice (Fig. 2a), we found that the values of EF% and FS% in 60-week-old aged mice were significantly lower than those in young 7-week-old mice, but this did not reach the condition of heart failure (Fig. 2b). There were no obvious Senescence-associated β-galactosidase (SA-β-gal)-positive cells in the heart tissue of 7-week-old mice, while the number of SA-β-gal positive cells in the heart tissue of 60-week-old mice was significantly increased (Fig. 2c). Western blotting showed that Rnd3 protein levels were decreased in the aged mice, accompanied by an increase in cellular senescence markers, such as p53, p16, Monocyte chemoattractant protein-1 (MCP1), interleukin (IL)-6, IL1-α, and Growth differentiation factor 15 (GDF15) (Fig. 2d-e).

**Figure 2.**
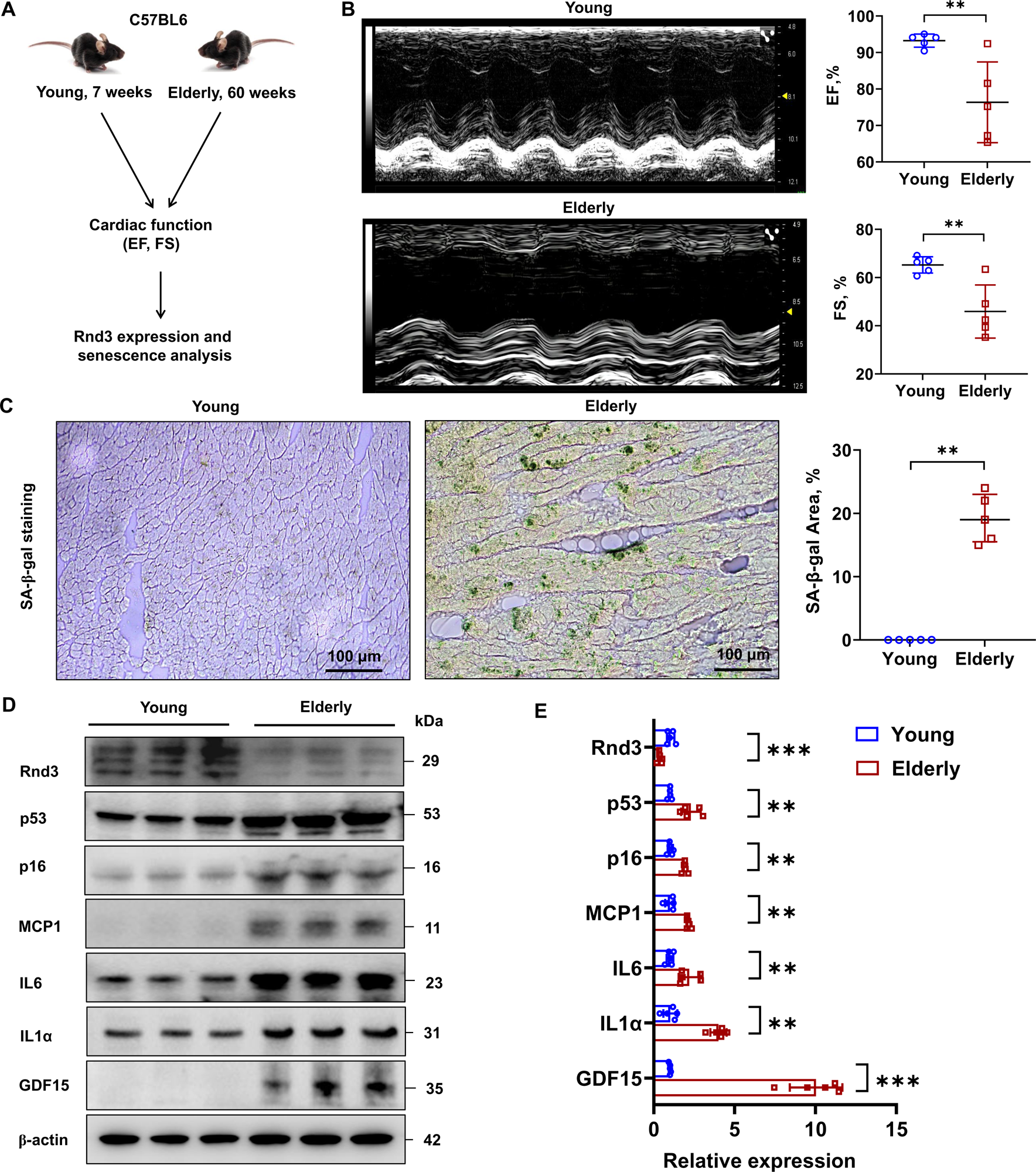
*Rnd3* expression and cardiac function in juvenile and aged mice. (**A**) Schematic diagram of the experimental procedure. (**B**) Ultrasound detection of cardiac function in young and older mice. Data were analyzed by the unpaired t test; \*\**P*<0.01. (**C**) SA-β-galactosidase staining using frozen sections of the mouse heart. Data were analyzed by the unpaired t test; \*\**P*<0.01. (**D**) Western blot detection of Rnd3 and cell cycle-related p53 and p16 protein levels, and the SASP-related proteins IL-1α, IL-6, and GDF15. ꞵ-actin served as an internal reference. (**E**) Quantitative analysis of Rnd3, p53, and p16 protein levels and the SASP-related genes IL-1α, IL6, and GDF15. Data were analyzed by the unpaired t test; \*\**P*<0.01, \*\*\**P*<0.001.

### Diabetes Inhibits *Rnd3* Expression in Heart Tissue and Induces Cardiomyocyte Senescence

In streptozotocin (STZ)-induced Type 1 diabetes mellitus (T1DM) rats and spontaneous Type 2 diabetes mellitus (T2DM) mice (Fig. 3a), Rnd3 expression in heart tissue was significantly lower than that in controls (Fig. 3b). Considering that cardiomyocytes highly expressed *Rnd3* in normal heart tissues (Fig. S1), we therefore consequently explore the role of diabetes on Rnd3 expression in cardiomyocytes. *In vitro* experiments also showed that high glucose (HG, 35 mmol/l D-glucose) stimulation effectively inhibited Rnd3 expression in cardiomyocytes (Fig. 3c), and this inhibition effect was glucose-dependent (Fig. S2). To further clarify the role of Rnd3 downregulation in cardiomyocyte senescence, we used CRISPR/Cas9 technology to construct H9C2 cells with *Rnd3* gene knockout (Fig. 3d). I*n vitro* HG stimulation significantly induced SA-β-gal-positive staining in H9C2 cells, and *Rnd3* gene knockout further amplified this effect (Fig. 3e). Additionally, DNA damage was aggravated (Fig. 3f) and the expression of SASP-related factors was upregulated (Fig. 3g, h).

**Figure 3.**
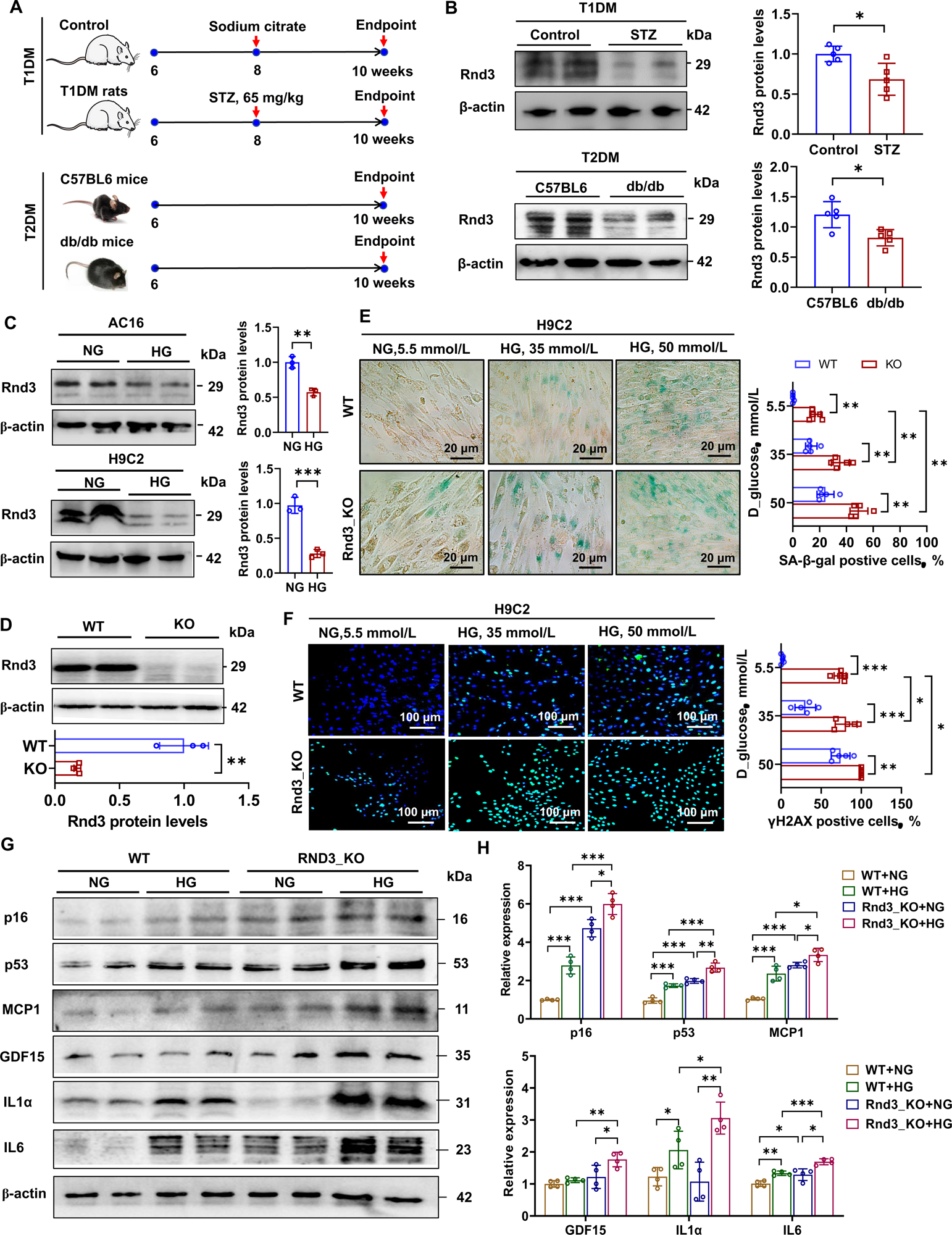
Effects of diabetes/high glucose on *Rnd3* expression and cell senescence in cardiomyocytes. (**A**) Schematic diagram of the establishment of T1DM and T2DM models. (**B**) Western blot detection of Rnd3 protein levels in heart tissue of diabetic rats and mice. ꞵ-actin served as an internal reference. Data were analyzed by the unpaired t test; \**P*<0.05. (**C**) Western blot detection of Rnd3 protein levels in high glucose (HG) (35 mmol/L D-glucose)-treated cardiomyocytes. Data were analyzed by the unpaired t test; \*\**P*<0.01, \*\*\**P*<0.001. (**D**) Establishment of *Rnd3* gene knockout H9C2 cardiomyocytes using CRISPR/Cas9 technology. Data were analyzed by the unpaired t test; \*\**P*<0.01. (**E**) SA-β-galactosidase staining in normal glucose (NG)- and HG-treated H9C2 cells. Data were analyzed by the unpaired t test; \*\**P*<0.01. (**F**) Immunofluorescence staining of γH2AX in NG (5.5 mmol/L D-glucose)-treated and HG (35 mmol/L D-glucose)-treated H9C2 cells. Data were analyzed by the unpaired t test; \**P*<0.05, \*\**P*<0.01, \*\*\**P*<0.001. (**G, H**) Western blot detection of cell cycle-related p53 and p16 protein levels, and the SASP-related proteins MCP1, IL-1α, IL-6, and GDF15 in NG (5.5 mmol/L D-glucose)-treated and HG (35 mmol/L D-glucose)-treated H9C2 cells. ꞵ-actin served as an internal reference. Data were analyzed by the unpaired t test, \**P*<0.05, \*\**P*<0.01, \*\*\**P*<0.001.

Subsequently, we constructed the T2DM rat model with cardiomyocyte-specific *Rnd3* gene knockout (KO) and overexpression (Fig. 4a. The characteristics of these rats are shown in Fig. S3, Table S2). Echocardiography showed that T2DM adversely affected ejection fraction, fractional shortening and the E/A ratio in the heart, and *Rnd3* KO further aggravated this damage. Restoring *Rnd3* expression delayed the damage of T2DM to heart function (Fig. 4b, c). Western blotting showed that *Rnd3* KO promoted in cellular senescence markers levels induced by T2DM in heart tissue, while overexpressing *Rnd3* reduced the production of cellular senescence markers in heart tissue (Fig. 4d, e). γH2AX and SA-β-gal staining showed that *Rnd3* KO promoted the accumulation of γH2AX- and SA-β-gal-positive cells in the heart tissue of diabetic rats, while *Rnd3* overexpression reversed this effect (Fig. 4f-i).

**Figure 4.**
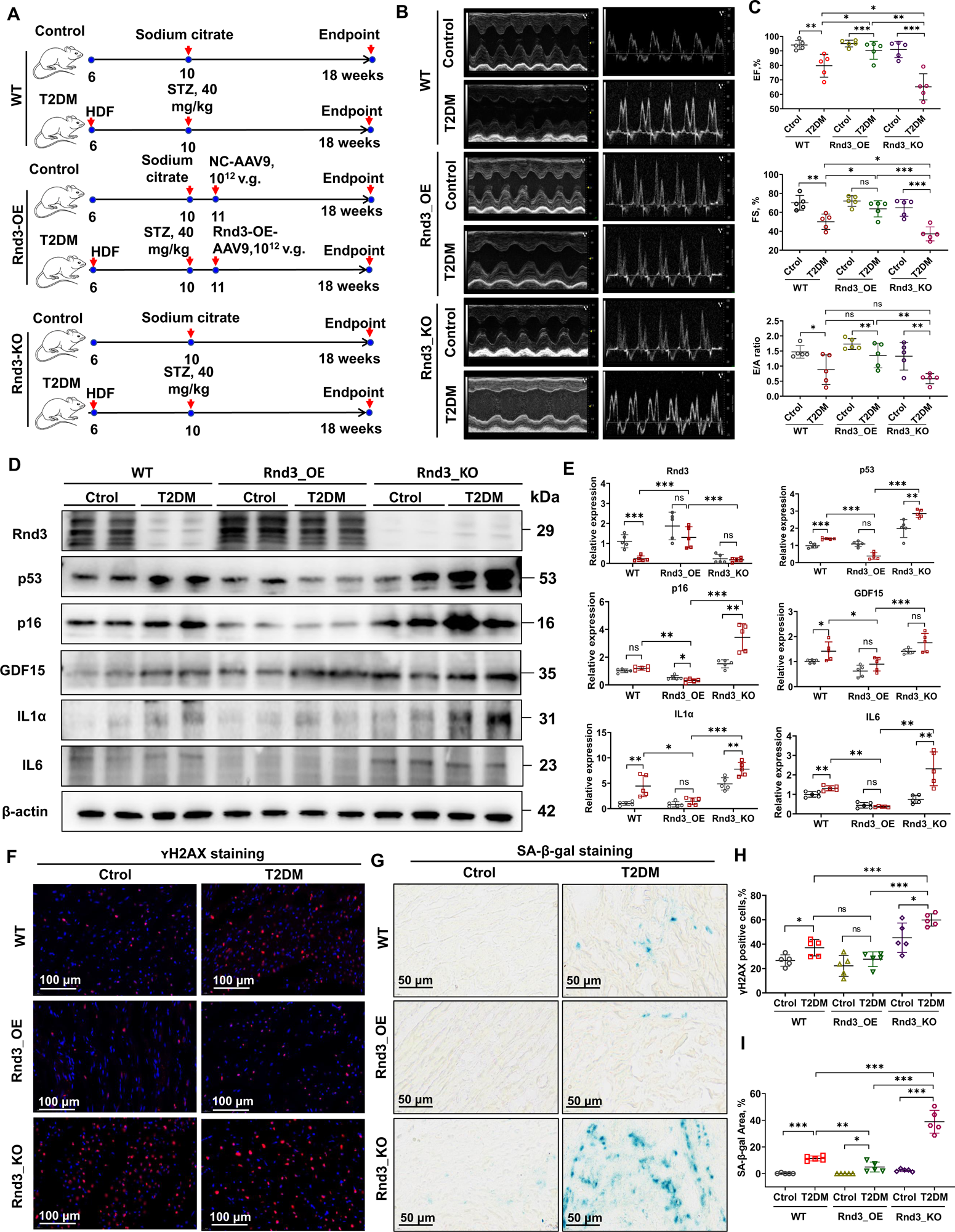
Cardiac function and cellular senescence analysis in rats with or without *Rnd3* gene-targeted intervention. (**A**) Schematic diagram of the establishment of T2DM rats. (**B**) Representative images of echocardiography. (**C**) Cardiac function in rats with or without *Rnd3* gene intervention. Data were analyzed by the unpaired t test; \**P*<0.05, \*\**P*<0.01, \*\*\**P*<0.001. ns, no significance. (**D**) Western blot detection of protein levels of Rnd3, cell cycle-related p53 and p16, and the SASP-related markers MCP1, IL-1α, IL-6, and GDF15 in rats with or without *Rnd3* gene intervention. ꞵ-actin served as an internal reference. (**E**) Quantitative analysis of protein levels of Rnd3, cell cycle-related p53 and p16, and the SASP-related factors MCP1, IL-1α, IL-6, and GDF15 in rats with or without *Rnd3* gene intervention. Data were analyzed by the unpaired t test, \**P*<0.05, \*\**P*<0.01, \*\*\**P*<0.001. ns, no significance. (**F, H**) Immunofluorescence staining of γH2AX in heart tissue of rats with or without *Rnd3* gene intervention. Data were analyzed by the unpaired t test; Bars = 100 μm. \**P*<0.05, \*\*\**P*<0.001. ns, no significance. (**G, I**) SA-β-galactosidase staining in frozen sections of the heart in rats with or without *Rnd3* gene intervention. Data were analyzed by the unpaired t test; Bars = 50 μm. \**P*<0.05, \*\**P*<0.01, \*\*\**P*<0.001, ns, no significance.

### HG Stimulation Upregulates miR-103a-3p to Inhibit *Rnd3* Expression in Cardiomyocytes

We hypothesized that microRNA (miRNA) plays a major role in the HG-induced inhibition of Rnd3 expression. To test this hypothesis, we performed miRNA sequencing on AC16 cells cultured in NG (5.5 mmol/L D-glucose) and HG (35 mmol/L D-glucose) to identify miRNAs that are regulated by glucose and target *Rnd3*. We found that, after 48 hours of treatment with HG, there were 43 differentially expressed miRNAs (23 upregulated and 20 downregulated) in AC16 cells compared with NG treatment (Table S3). Considering that most miRNAs inhibit the expression of their target genes. Therefore, we focused on miRNAs that were upregulated by HG stimulation. We observed that miR-103a-3p was closely related to T2DM and cellular senescence (Fig. 5a). Further experiments showed that miR-103a-3p expression was increased in cardiomyocytes stimulated by HG *in vitro* (Fig. 5b). By comparing the base sequences of miR-103a-3p and *Rnd3*, we found that miR-103a-3p was highly conserved between humans and rats. There was one binding site at position 1379 on the 3′UTR of the human *Rnd3* gene, and two binding sites at positions 579 and 716 on the 3′UTR of the rat *Rnd3* gene (Fig. 5c). The results of the luciferase reporter gene experiment supported the inhibitory effect of miR-103a-3p on the 3′UTR promoter of the *Rnd3* gene (Fig. 5d). We also examined miR-103a-3p concentrations in plasma of a clinical cohort including 24 cases of T2DM and 17 controls (see Table S4 for the clinical data of individuals). Interestingly, miR-103a-3p concentrations were significantly higher in the clinical T2DM population than in the non-diabetic controls (Fig. 5e). Additionally, miR-103a-3p concentrations were positively correlated with FBG concentrations (r=0.408, *P*=0.0041) and the triglyceride and glucose index (r=0.441, *P*=0.002) (Fig. 5f).

**Figure 5.**
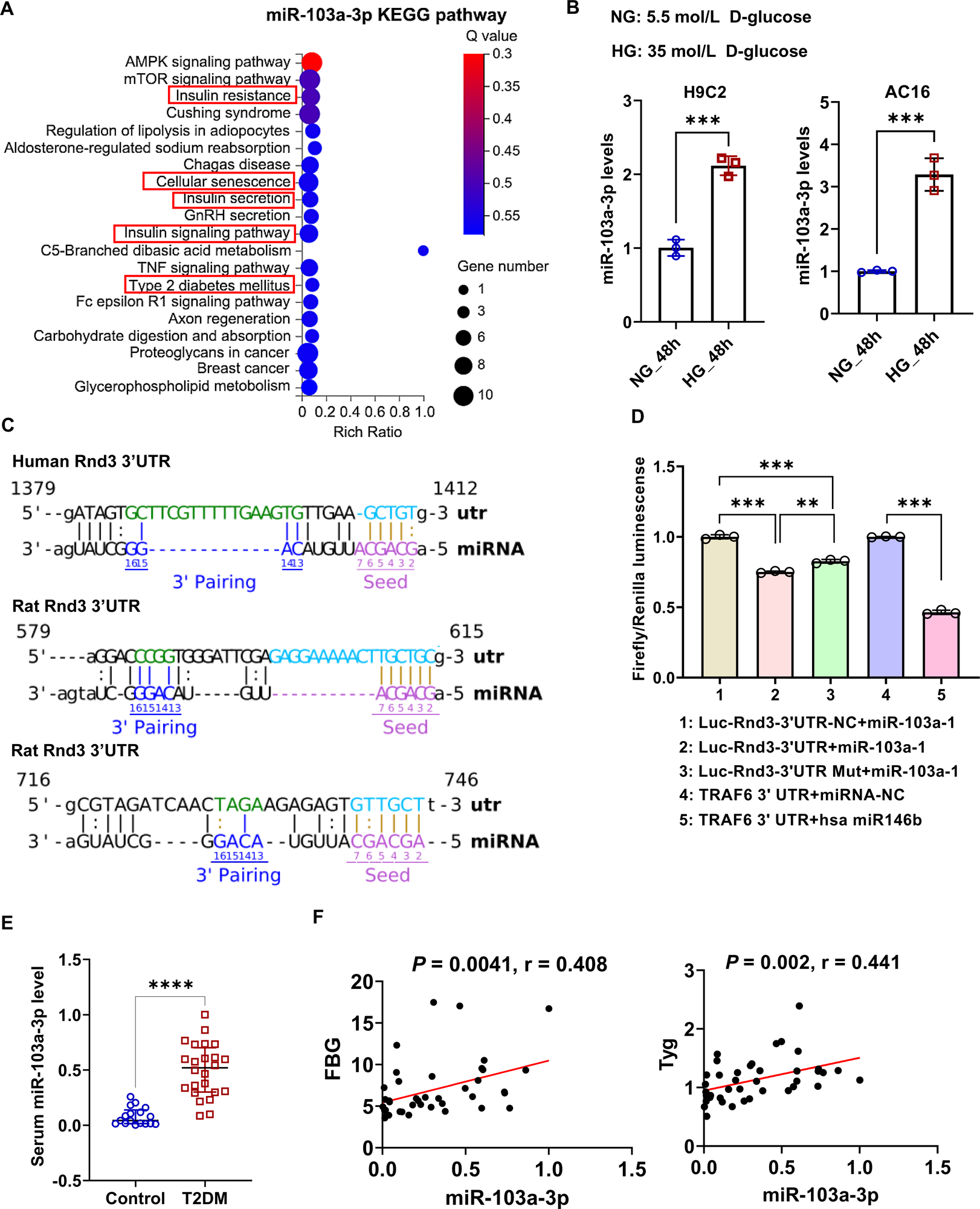
Function and expression of miR-103a-3p. (**A**) Kyoto Encyclopedia of Genes and Genomes (KEGG) analysis of miR-103a-3p. KEGG items in the red box indicate that miR-103a-3p is related to cellular senescence and diabetes. (**B**) Quantitative RT-PCR detection of miR-103a-3p in NG (5.5 mmol/L D-glucose, 72 hours)-treated and HG (35 mmol/L D-glucose, 72 hours)-treated cardiomyocytes. Data were analyzed by the unpaired t test; \*\*\**P*<0.001. (**C**) Analysis of binding sites between miR-103a-3p and the human and rat *Rnd3* gene 3′UTR. (**D**) The dual luciferase reporter gene method was used to detect the binding effect of miR-103a-3p with the rat *Rnd3* gene 3′UTR. Groups 4 and 5 served as negative and positive controls to demonstrate the reliability of the experimental system. Data were analyzed by the Bonferroni post-hoc test post-one-way analysis of variance; \*\*\**P*<0.001. (**E**) Quantitative RT-PCR detection of miR-103a-3p in serum samples of a clinical cohort of T2DM (*n*=24) and controls (*n*=17). Data were analyzed by the Mann–Whitney test; \*\*\*\**P*<0.0001. (**F**) Pearson correlation analysis of miR-103a-3p levels with fasting blood glucose (FBG) and the triglyceride and glucose index (Tyg) in a clinical cohort (*n*=41).

### Downregulating miR-103a-3p *in vitro* Restores Rnd3 Expression and Alleviates HG-Induced Cardiomyocyte Aging

To investigate the therapeutic value of targeting miR-103a-3p for diabetes-induced cardiomyocyte aging, we designed an inhibitor molecule for miR-103a-3p and applied it to H9C2 cardiomyocytes stimulated with HG (35 mmol/L D-glucose). After the administration of the miR-103a-3p inhibitor, *Rnd3* expression levels in cardiomyocytes were significantly increased, accompanied by downregulation of cellular senescence related genes expression (Fig. 6a, b, Fig. S4). SA-β-gal and γH2AX staining suggested that the miR-103a-3p inhibitor effectively alleviated diabetes-mediated cardiomyocyte senescence and DNA damage (Fig. 6c, d). These findings laid the foundation for the following *in vivo* experiments.

**Figure 6.**
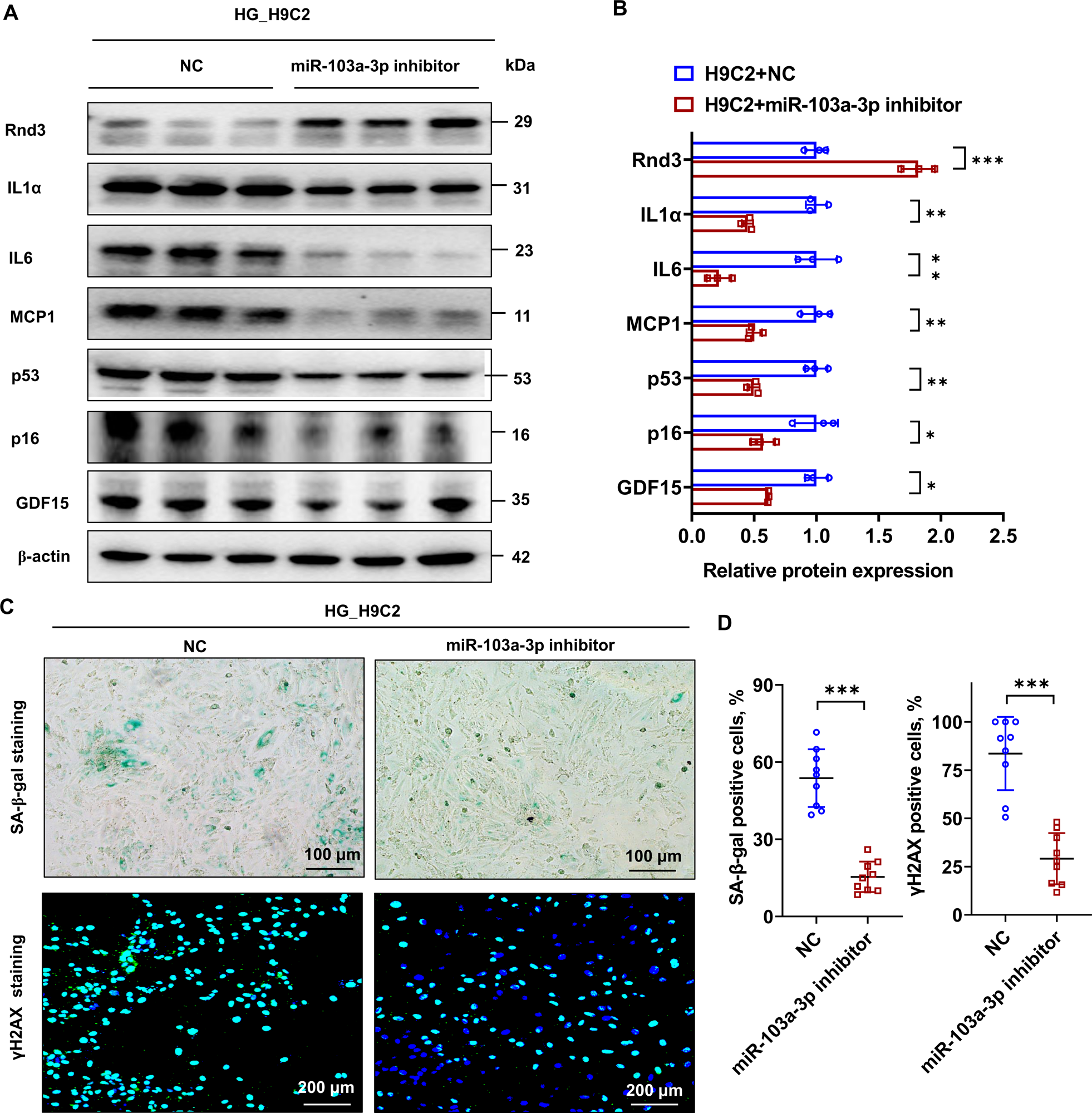
Effects of an miR-103a-3p inhibitor on *Rnd3* expression and cardiomyocyte senescence. (**A**) Western blot detection of Rnd3 and cell cycle-related p53 and p16 protein levels, and the SASP-related markers MCP1, IL-1α, IL-6, and GDF15 in HG (35 mmol/L)-treated H9C2 cells. ꞵ-actin served as an internal reference. (**B**) Quantitative analysis of protein levels of Rnd3, cell cycle-related p53 and p16, and the SASP-related markers MCP1, IL-1α, IL-6, and GDF15 in HG (35 mmol/L)-treated H9C2 cells. Data were analyzed by the unpaired t test; \**P*<0.05, \*\**P*<0.01, \*\*\**P*<0.001. (**C**) SA-β-galactosidase and γH2AX staining in HG (35 mmol/L)-treated H9C2 cells. Data were analyzed by the unpaired t test; \*\*\**P*<0.001.

### Cardiac Overexpression of miR-103a-3p Sponges Restores *Rnd3* Expression and Improves Heart Function in Diabetic Rats

To further evaluate the therapeutic effect of antagonizing miR-103a-3p on diabetes-induced myocardial aging, we constructed a T2DM rat model by a high-fat diet supplemented with intraperitoneal injection of STZ. One week later, AAV9-overexpressing miR103a-3p_sponges and negative control AAV9 were infused into the rat tail vein (Fig. 7a. The characteristics of these rats are shown in Table S5). We found that the EF%, FS% and E/A peak values in diabetic rats in the overexpressed miR-103a-3p_sponge group were significantly better than those in diabetic rats injected with the control virus (Fig. 7b, c). Western blotting showed that overexpression of miR103a-3p_sponges effectively relieved the inhibitory effect of T2DM on Rnd3 protein levels in cardiac tissue, and also reduced SASP protein levels (Fig. 7d, e), consistent with the results of PCR (Fig. S5). The positive rates of SA-β-gal and γH2AX staining in myocardial cells in T2DM rats were significantly higher than those in controls. Additionally, the positive rates of SA-β-gal and γH2AX staining in T2DM rats infused with AAV9-miR-103a-3p_sponges were significantly lower than those in controls (Fig. 7e-h). These findings suggest that inhibiting miR-103a-3p and thereby restoring *Rnd3* expression is an effective means of disrupting diabetes-induced myocardial aging.

**Figure 7.**
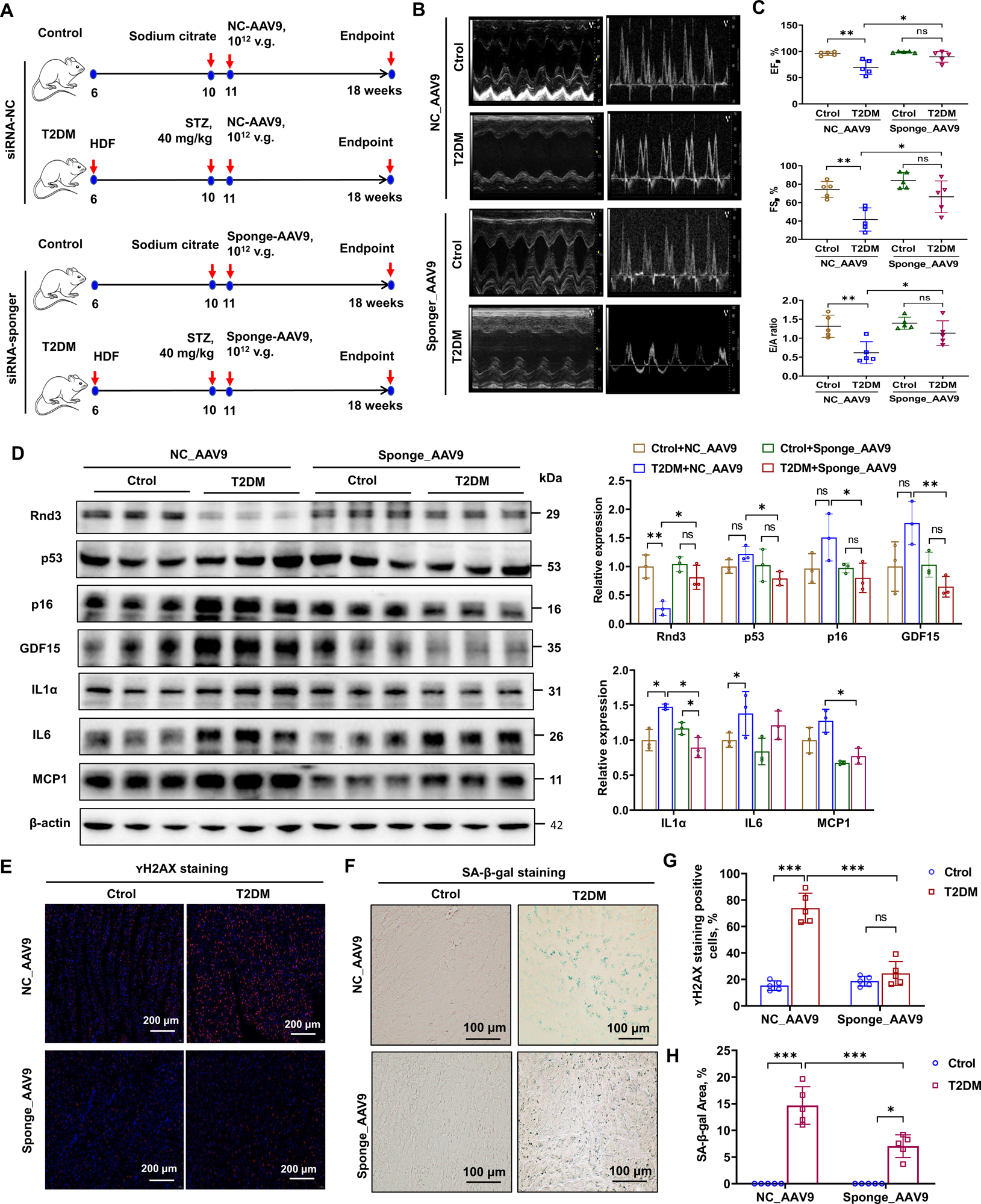
Cardiac function and cellular senescence analysis in rats treated with miR-103a-3p sponges. (**A**) Schematic diagram of the establishment of T2DM rats. (**B**) Representative images of echocardiography. (**C**) Cardiac function in rats with or without miR-103a-3p inhibition. Data were analyzed by the unpaired t test; \**P*<0.05, \*\**P*<0.01. ns, no significance. (**D**) Western blot detection of Rnd3 and cell cycle-related p53 and p16 protein levels, and the SASP-related markers MCP1, IL-1α, IL-6, and GDF15 in rats with or without miR-103a-3p inhibition. ꞵ-actin served as an internal reference. Data were analyzed by the unpaired t test; \**P*<0.05, \*\**P*<0.01. ns, no significance. (**E, G**) Immunofluorescence staining and quantitative analysis of γH2AX in heart tissue of rats with or without miR-103a-3p inhibition. Data were analyzed by the unpaired t test; \*\*\**P*<0.001. ns, no significance. (**F, H**) SA-β-galactosidase staining in frozen sections of the heart in rats with or without miR-103a-3p inhibition. Data were analyzed by the unpaired t test; \**P*<0.01, \*\*\**P*<0.001. ns, no significance.

### Downregulation of Rnd3 Induces a Cellular Senescence Phenotype in Cardiomyocytes by Activating the STAT3 Pathway

Finally, we examined the potential mechanism by which HG-suppressed Rnd3 expression promotes cardiac aging. STAT3, which is a transcription factor related to inflammation-associated SASP, may mediate this effect. T2DM rats showed higher phosphorylated (p)-STAT3(Tyr705) levels than control wild type rats, but Rnd3 overexpression was inhibited, while *Rnd3* knockout enhanced p-STAT3 (Tyr705) levels in the T2DM rat heart (Fig. S6a). Furthermore, *in vitro* HG stimulation led to the upregulation of p-STAT3(Tyr705) in cardiomyocytes, and *Rnd3* knockout amplified the level of p-STAT3 (Tyr705) stimulated by HG (Fig. S6b). While administering S3I-201, p-STAT3 (Tyr705) levels decreased, and the expression of all proteins related to SASP (GDF15, IL-1α, IL-6, and MCP1), except for p53 and Rnd3, was also reduced (Fig. 8a, Fig. S6c). We also confirmed that HG stimuli could profoundly activated p-STAT3 (Tyr705) in *Rnd3* KO cardiomyocytes (Fig. 8b). Mechanistically, *Rnd3* KO led to obviously attenuated p-STAT3(Tyr705) ubiquitination through proteasome (Fig. 8c). what’s more, a binding between Rnd3 and total STAT3 in cardiomycytes was detected (Fig. 8d). Interestingly, S3I-201 attenuated HG stimuli-induced nuclei accumulation of p-STAT3(Tyr705) (Fig. 8e), as well as HG-induced SA-β-gal-positive cells mostly in *Rnd3* knockout cells (Fig. 8f). These findings suggest that the STAT3 signal is involved in cardiac cell aging induced by the downregulation of Rnd3 expression and is mediated by diabetic stimulation. Overall, the activation of the miR-103a-3p/Rnd3/STAT3 signal plays an important role in diabetes-induced cardiac cell aging, and also has the potential for targeted intervention (Fig. 8g).

**Figure 8.**
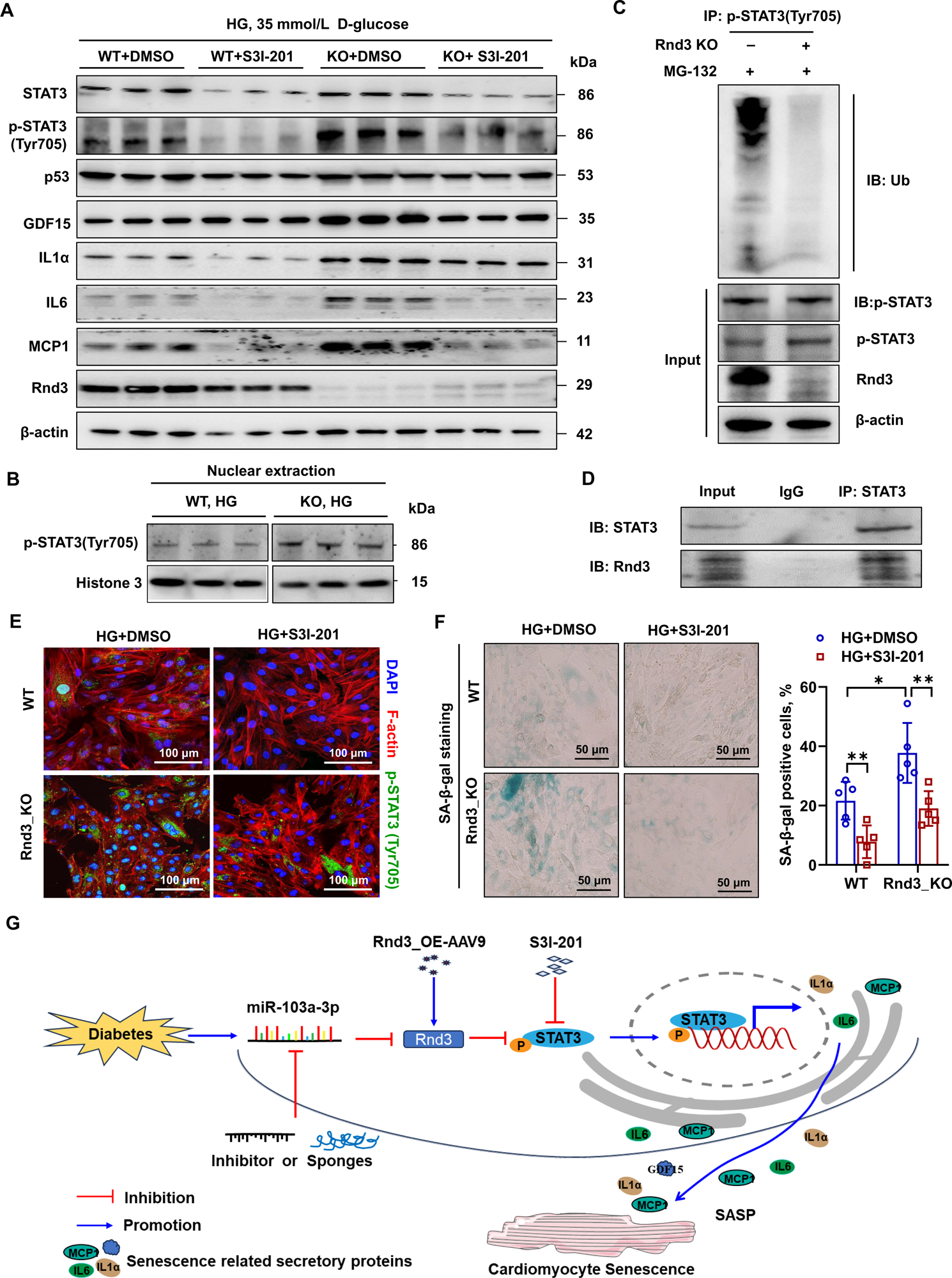
Interaction of Rnd3 with STAT3 in cardiomyocytes and its role in high glucose (HG) -induced cellular senescence. (**A**) Western blot detection of the STAT3 inhibitor S3I-201 and its effect on STAT3 activation and cellular senescence in HG (35 mmol/L D-glucose) -treated H9C2 cells. ꞵ-actin served as an internal reference. (**B**) Upregulation of p-STAT3(Tyr705) in H9C2 cells post HG (35 mmol/L D-glucose) stimuli for 48h. Histone 3 served as an internal reference. (**C**) Ubiquitination mediated p-STAT3(Tyr705) degradation in wild and *Rnd3* KO H9C2 cardiomyocytes. (**D**) Interaction of Rnd3 and STAT3 assessed by Co-Immunoprecipitation. (**E**) Immunofluorescence detection of the expression and intracellular localization of p-STAT3(Tyr705) in HG (35 mmol/l D-glucose) stimulated H9C2 cells by STAT3 inhibitor S3I-201. Bars = 100 μm. (**F**) STAT3 inhibitor S3I-201 suppresses HG (35 mmol/l D-glucose) induced H9C2 cardiomyocyte senescence. Bars = 50 μm. \**P*<0.05, \*\**P*<0.01. (F)Schematic diagram of the mechanism of how diabetes mediates cardiomyocyte senescence. Diabetes upregulates miR103a-3p expression, further inhibiting Rnd3 expression in cardiomyocytes, and then activates STAT3 signaling to induce the SASP phenotype, and finally mediates the senescence of cardiomyocytes. An miR-103a-3p inhibitor, STAT3 inhibitor, and Rnd3-targeted recovery by AAV9 are helpful to treat cardiomyocyte senescence induced by diabetes.

## DISCUSSION

Rnd3 has become the focus of attention from scholars worldwide for its association with cancer.^17^ Our research group, in collaboration with Dr Chang, has been investigating the relationship between *Rnd3* and cardiovascular disease for more than a decade.^17,19,21,22^ Other research groups have also suggested that *Rnd3* plays a crucial role in vascular and cardiac remodeling.^23,24^ These findings suggest that *Rnd3* may have considerable therapeutic value in cardiovascular disease. This study further extends the role of *Rnd3* in diabetes-induced cardiomyocyte aging. We found that downregulation of *Rnd3* expression exacerbated diabetes-induced cardiomyocyte senescence, possibly due to diabetes-induced elevation of miR-103a-3p, which directly inhibits *Rnd3*. This process then activated STAT3 signaling and promoted cardiomyocyte senescence. Interestingly, targeted inhibition of miR-103a-3p and overexpression of *Rnd3* helped in restoring *Rnd3* expression in DCM and reversed cardiomyocyte senescence. This study provides a potentially feasible strategy for the prevention and treatment of DCM aging.

Since the Framingham study first reported that diabetes can aggravate cardiovascular mortality,^25^ the rapidly growing number of diabetes populations worldwide has led to attention being paid to the cardiovascular complications of diabetes. Our study first examined the relationship between *Rnd3* and diabetes using the GEO dataset, and we found downregulation of *Rnd3* mRNA levels in patients with diabetes. According to the International Diabetes Federation, the global population with diabetes was approximately 463 million in 2019 and is expected to increase to 578 million by 2030.^26^ Cardiovascular events associated with diabetes have become major causes of death and disability in diabetes, making the prevention and treatment of diabetes a major global public health issue. These results based on the GEO public database provide a platform for further in-depth exploration of the relationship and regulatory mechanism between abnormal *Rnd3* expression and diabetes.

The long-term pathological state of diabetes, such as hyperglycemia, hyperlipidemia, insulin resistance, and oxidative stress, can accelerate cardiac cell aging and injury.^8,27,28^ This process is different from the mechanism of cell aging caused by a simple increase in age. In our cohort, we found that *Rnd3* mRNA levels were negatively correlated with the population’s age (i.e., as age increased, *Rnd3* mRNA levels decreased), suggesting a relationship between *Rnd3* levels and aging. This result is consistent with the results of Rnd3 expression, heart function, and cellular senescence markers in the heart tissue of young and old mice. Further correlation analysis showed that *Rnd3* levels were also related to some factors of myocardial injury, such as cTnI, myoglubin, NT-proBNP and cardiac function markers like EF% and E/A ratio. These results further confirm our initial assumption. Therefore, we will verify our results through cell and animal experiments in subsequent research.

The approach based on single-cell RNA sequencing has recently received much attention, providing the possibility to analyze the composition of organ and cell changes under different states.^29^ Using bioinformatics, we found that normal human heart tissue contained various cell types, and the *Rnd3* gene had the highest abundance of expression in certain subtype of cardiomyocytes. These findings suggest that *Rnd3* plays a crucial role in cardiac cells. On the basis of this finding, we constructed a rodent model of cardiac *Rnd3* overexpression and knockout, and obtained H9C2 cardiomyocytes with stable knockout of the *Rnd3* gene using CRISPR/Cas9 technology. The successful establishment of these animal and cell models laid a solid foundation for the implementation of this study.

On the basis of the finding that *Rnd3* expression was present in diabetic heart tissue and HG-treated cardiomyocytes, we constructed H9C2 cells with *Rnd3* gene knockout based on previous protocols.^30^ Our study showed that *Rnd3* knockout cells stimulated by HG were more likely to show cell cycle arrest and SASP. SASP is a key feature of cell aging, and related factors play a decisive regulatory role in processes, such as cellular senescence, inflammation, immune modulation, growth arrest, fibrosis, and tumorigenesis.^31^ Therefore, the lack of *Rnd3* expression contributes to HG-induced cardiomyocyte senescence. Indeed, *Rnd3* gene knockout amplified the diabetes-induced myocardial damage. In contrast, overexpressing *Rnd3* in heart tissue through tail vein injection of AAV9 restored heart function and alleviated the SASP. Considering the clinically significant relevance of cellular senescence in the management of patients with cardiac diseases,^32^ the current study suggests that targeting *Rnd3* to reverse myocardial cell aging could considerably enhance the treatment of DCM.

Upon finding that diabetes induced cardiomyocyte senescence by suppressing *Rnd3* expression, this study focused on the epigenetics of miRNA. The miRNAs are endogenous non-coding RNAs of approximately 22 nt in length, and they play a universal inhibitory role in gene silencing.^33^ Ihe potential of miRNA as a therapeutic target for cardiovascular aging is gradually gaining attention.^34,35^ We hypothesized that there are certain miRNAs that target the *Rnd3*′UTR and they are upregulated during diabetes. To test this hypothesis, we performed miRNA sequencing on AC16 cells cultured under different glucose concentrations to screen for differentially expressed miRNAs. We discovered numerous differentially expressed miRNAs in AC16 cardiomyocytes stimulated by HG. We searched for the conservation of differentially expressed miRNAs among species through the microRNA viewer website, and prioritized miR-103a-3p, which is fully conserved between humans and rats, for further research. Bioinformatics analysis showed that the target genes of miR-103a-3p were involved in numerous pathological physiological processes, such as aging, cardiovascular disease, and endocrine disease. This miRNA plays a regulatory role in various biological processes, such as sugar, lipid, vitamin, and energy metabolism, tissue regeneration, signal transduction, and inflammatory responses. Using quantitative RT-PCR, we found that miR-103a-3p expression in human and rat cardiomyocytes was increased under glucose stimulation, suggesting that miR-103a-3p may be an appropriate target in diabetic damage. Subsequent *in vitro* experiments suggested that directly using an miR-103a-3p inhibitor restored *Rnd3* expression in H9C2 cells under HG conditions, and simultaneously reduced HG-induced cardiomyocyte senescence. Using *in vivo* experiments, our study further showed that overexpressing miR-103a-3p sponges delayed the damage in heart function caused by diabetes in rats, and inhibited the expression of cellular senescence related factors in rat heart tissue. Most importantly, we found that blood glucose concentration-related circulating miR-103a-3p levels were upregulated in the diabetes population compared with non-diabetes controls. Therefore, miR-103a-3p could be involved in diabetes-suppressed *Rnd3* expression. No gross complications were observed in rats administered miR-103a-3p sponges, and their survival rate was not different from that in the control group during the experimental period. However, the safety of AAV9-administered miR-103a-3p sponges in other tissues and organs still needs more comprehensive verification. There are more than 150 reports regarding miR-103a-3p in PubMed; among them, only one report focused on cardiomyocytes,^36^ and one meta-analysis suggested that miR-103a-3p could be a biomarker for T2DM.^37^ Our current research expands the application of miR-103a-3p in DCM treatment.

Rnd3 reduces oxidative stress, cell proliferation, fibrosis, and the inflammatory response by inhibiting ROCK1, nuclear factor kappa-B, Notch, and transforming growth factor-β signals in various cells and tissues,^18,22,23,34^ and these signals are also crucial for driving cardiac aging. Our previous study showed that *Rnd3* knockout activated the Jak/STAT pathway.^30^ Activation of STAT3 signaling is pivotal for cell-to-cell communication in the heart.^38^ This finding laid the foundation for our subsequent analysis of how Rnd3 downregulation mediates cardiomyocyte senescence. Reports have shown that p-STAT3 is upregulated with HG stimulation and promotes the expression of SASP-related inflammatory factors,^39,40^ as found in this study. Most important, our current investigation for the first time revealed that Rnd3 could interacts with STAT3 in cardiomyocyes, and this binding provoked the ubiquitination of p-STAT3. These findings suggest that HG stimulation and *Rnd3* knockout upregulate p-STAT3 expression, with its mechanism was mostly related to ubiquitination mediated degradation pathway.

STAT3 activation plays protective or pathogenic roles under different pathophysiological conditions in the cardiovascular system. IL-6 is a key activator of STAT3. IL-6 mediates left ventricular hypertrophy and myocardial fibrosis by activating STAT3 signaling.^41,42^ However, Granulocyte Colony Stimulating Factor prevents post-infarction ventricular remodeling by activating the JAK/STAT pathway.^43^ Moreover, cardiac-specific knockout of STAT3 exacerbates subacute ventricular remodeling in mice with myocardial infarction.^44^ In the current study, we observed that the expression of SASP-related factors, such as IL-6, MCP1, and IL-1α, was increased with the knockout of *Rnd3*, along with upregulation of p-STAT3. These SASPs and the STAT3 pathway have a mutual regulatory effect. In particular, IL-6 and MCP1 can promote STAT3 phosphorylation, and STAT3 phosphorylation further increases the expression levels of IL-6 and MCP1.^41,45,46^ To further clarify the upstream and downstream relationship between Rnd3, SASP, and STAT3, we used the highly selective STAT3 inhibitor S3I-201^47^. Interestingly, regardless of the group (wild type or *Rnd3* knockout), treating H9C2 cells with S3I-201 significantly inhibited the expression of SASP-related factors, such as IL-1α, IL-6, and MCP1, but not Rnd3, in parallel with the attenuation of SA-β-gal-positive cells. These findings suggest that the activation of STAT3 is a key pathway involved in diabetes-/HG-induced cardiomyocyte senescence through inhibiting *Rnd3*. A recent reported showed that blocking aged-related inflammatory responses is important in the treatment of aging and degeneration of the nervous system.^48^ Our study showed that targeted intervention of miR-103a-3p/Rnd3/STAT3 signaling in diabetes blocked the expression of SASP-related inflammatory factors and thus reduced cardiomyocyte cell aging, which is consistent with the above-mentioned findings.

## LIMITATIONS

There are some limitations to our study. First, the heart is composed of various cell types. Our study only focused on the effect of *Rnd3* expression defects under diabetes on the aging of cardiomyocytes. Further research is required to investigate whether there are other mechanisms between various cells that affect cardiac aging. Second, abnormal *Rnd3* expression also involves other signaling pathways that affect myocardial aging, Therefore, using multiomics such as proteomics methods may reveal more signaling pathways affected by *Rnd3* dysregulation.

## CONCLUSION

Diabetes induces cardiomyocyte senescence by upregulating miR-103a-3p, thereby suppressing *Rnd3* expression and subsequently activating STAT3 to elevate the transcription levels of SASP-related factors. Direct myocardial overexpression of *Rnd3* or genetic inhibition of miR-103a-3p or pharmacological inhibition of STAT3 can preserve cardiac function in diabetic rats and counteract cardiomyocyte cell senescence.

## Affiliations

Hainan Provincial Key Laboratory for Tropical Cardiovascular Disease Research, the First Affiliated Hospital, Hainan Medical University, Haikou, China (L. Wu, C. Luo, X. Zhu, Y. Chen, Y. Liu, Y. Zhao, S. Pan, K, Shi, X. Ling, J. Qiu, S. Zhang, J. Chai, J. Guo, W. Jie), Department of Pathology and Pathophysiology, School of Basic Medicine Sciences, Guangdong Medical University, Zhanjiang, China (J. Xing, Z. Shen), and Department of Medicine, Brigham and Women’s Hospital of Harvard Medical School, Boston, MA, USA (X. Ling).

## Author Contributions

L. Wu, X. Zhu, C. Luo, Y. Chen, Y. Zhao, S. Pan, J. Xing, K. Shi, J. Qiu and S. Zhang performed the experiments and collected the data. Y. Liu and X. Ling collected the clinical samples and reviewed the data. L. Wu, XLZ, CL, and JXC prepared the materials. Z. Shen and X. Ling searched the literature and performed statistical analyses. Z. Shen, J. Guo, and W. Jie conceived and coordinated the study and drafted the manuscript. All of the authors read and approved the final manuscript.

## Data availability statement

The original contributions presented in the study are included in the article/Supplemental Material. Further inquiries can be directed to the corresponding authors.

## Ethics statement

All animal experiments conform to the ethical requirements for experimental animals of Hainan Medical University (Approval number: KYLL-2020-048). The use of human tissues was reviewed and approved by the Ethics Committee of the First Affiliated Hospital of Hainan Medical University (Approval number: 2023-KYL-096).

## Novelty and Significance

### What Is Known?

1. Rnd3 plays an important role in the setting of certain of cardiovascular diseases, including inflammatory responses after myocardial infarction, diabetic myocardial fibers, pressure overload-induced cardiomyocyte apoptosis, angiogenesis after heart failure, and arrhythmia.
2. Knockout of Rnd3 in cardiomyocytes leads to activation of JAK/STAT signaling, and this activation plays a protective or pathogenic role in heart disease.
3. Dysregulation of miR-103a-3p contributes to cancer progression, venous thromboembolism, and organic inflammation, fibrosis, and injury.

### What New Information Does This Article Contribute?

1. Rnd3 expression in myocardium was repressed by diabetes and aging.
2. Rnd3 deficiency synergistically contributed to diabetes-induced cardiomyocyte senescence, and overexpression of Rnd3 reversed diabetes-induced cardiomyocyte senescence.
3. Diabetes-induced miR-103a-3p directly bound *Rnd3* 3′UTR and thus modulated *Rnd3* expression, which participated in diabetes-induced cardiomyocyte senescence.
4. Rnd3 promote ubiquitination mediated degradation of STAT3. Activation of STAT3 was involved in *Rnd3* knockout enhancing diabetes-induced cardiomyocyte senescence. Our findings suggest that diabetes upregulates miR-103a-3p to inhibit Rnd3 expression, thereby promoting cardiomyocyte senescence, DNA damage, and cardiac dysfunction, and this is mediated by activation of the STAT3 pathway. Targeted inhibition of miR-103a-3p and upregulation of *Rnd3* expression can alleviate the pathological phenotype of diabetes-induced cardiomyocyte senescence, thereby delaying the progression of diabetic cardiomyopathy.

## Nonstandard Abbreviations and Acronyms

DCM: Diabetes cardiomyopathy
cTnI: Cardiac troponin I
E/A: Ratio of E/A
GDF-15: Growth differentiation factor 15
HG: High glucose
IL1a: Interleukin-1α
IL6: Interleukin-6
MCP-1: Monocyte chemoattractant protein-1
NG: Normal glucose
SASP: Senescence-associated secretory phenotype
STAT3: Signal transducer and activator of transcription 3
STZ: Streptozocin
T1DM: Type 1 diabetes mellitus
T2DM: Type 2 diabetes mellitus

