## Supplemental Materials and Methods and Figures,Tables for "Diabetes Advances Cardiomyocyte Senescence through miR-103a-3p/Rnd3/STAT3 Signaling"

#### Bioinformatics analysis based on the Gene Expression Omnibus and the Human Protein Atlas databases

The normalized probe expression matrices of diabetes were obtained from the Gene Expression Omnibus (GEO)(https://www.ncbi.nlm.nih.gov/gds/) with accession numbers: GSE23561, GSE95849, and GSE13760, respectively. Subsequently, the probe ID was converted into a gene symbol, and batch effects were eliminated from different datasets using the remove Batch Effect function of the limma(v3.52.4) . *Rnd3* gene expression in various cell types in cardiac tissue was analyzed using the Human Protein Atlas (https://www.proteinatlas.org/). The original sequencing data that had already been quantified was downloaded, and different cell types were annotated on the basis of known tissue and cell-specific markers. The primary cell types were selected to calculate and compare their *Rnd3* expression levels.

#### Animal models

Male C57BL/6 mice of 7 weeks and 60 weeks of age, as well as 8-week-old db/db mice, were procured from the Shanghai Model Organisms Center, Inc. (Shanghai, China). Male C57BL/6 mice of varying ages were used to simulate age-related senescence, while db/db mice were used to replicate spontaneous type 2 diabetes mellitus (T2DM). Five-week-old male wild-type Sprague–Dawley (SD) rats were purchased from Slaccas Company (Changsha, China) and were injected with streptozotocin (STZ) to induce the type 1 diabetes mellitus (T1DM) and T2DM models. Cardiomyocyte-specific *Rnd3* knockout mice were obtained through cross-breeding Flop+/+ rats and α-Myosin heavy chain(αMHC-Cre) rats (Cyagen, Suzhou, China). To establish the T1DM model, wild-type SD rats were intraperitoneally injected with 65 mg/kg STZ (#60256ES80; Yeasen, Shanghai, China). To establish the T2DM rat model, wild-type and cardiomyocyte *Rnd3* knockout SD rats were first fed a high-fat diet for 1 month (#PSY1102; Slaccas Company), followed by an intraperitoneal injection of 40 mg/kg STZ. Control rats were fed a normal diet and injected intraperitoneally with an equivalent volume of citrate buffer.

Blood

glucose concentrations were regularly measured, with fasting blood glucose concentrations ≥16.7 mmol/L indicating successful modeling. Some T2DM rats had tail vein injection of overexpressed miR_103a-3p sponges or control adeno-associated virus serotype 9(AAV9)($10^{12}$v,g/rat; Genechem, Shanghai, China). At the end point of the experiment, cardiac function including EF, FS, E/A of the animals was analyzed through echocardiography using Vevo 3100LT system (Fujifilm Visualsonics Inc., Toronto, ON, Canada), and heart tissue was used for related experiments.

**Human blood samples and clinical data**

From 1 January 2023 to 30 November 2023, patients treated at the Department of Cardiology of the First Affiliated Hospital of Hainan Medical University were selected according to the inclusion and exclusion criteria. Finally, 74 individuals including 42 T2DM and 32 normal controls were included in the study. Peripheral blood samples were collected, and serum and mononuclear cells were used for the extraction of RNA. The clinical data of each individual were also collected.

**Cell culture and treatment**

H9C2 rat cardiomyocytes were procured from the Shanghai Stem Cell Bank of the Chinese Academy of Sciences, while human cardiomyocytes (AC16 cells) were acquired from ATCC (Manassas, VA, USA). H9C2 and AC16 cells were maintained in Dulbecco's modified Eagle's medium supplemented with 10% fetal bovine serum, and the cells were incubated in an environment with 5% $CO_2$ at a saturated humidity of 37ºC. To examine the effect of varying glucose concentrations on cellular senescence, cells were cultured in Dulbecco's modified Eagle's medium with final concentrations of 12.5, 35, and 50 mmol/L D-glucose (high glucose [HG]), using a medium with a final concentration of 5.5 mmol/L D-glucose as a control (normal glucose [NG]). Cells at different time points were used for relevant experiments. To investigate the effect of miR103a-3p, H9C2 cells treated with 35 mmol/L D-glucose were subjected to treatment with an miR-103a-3p inhibitor (10 μmol/L; Ruibo, Guangzhou, China) for 48 hours. To examine the role of signal transducer and activator of transcription 3 (STAT3) signaling in HG-stimulated cardiomyocytes, the STAT3 inhibitor S3I-201 (10 μmol/L; MCE China, Shanghai) was used to stimulate the cells.

**Constructing *Rnd3* gene knockout in H9C2 cells**

*Rnd3* gene knockout H9C2 cells were established according to previous protocols [1]. Puromycin-stressed lentivirus infected cells were used for relative experiments.

**Senescence-associated -β-galactosidase staining**

Cells cultured in a six-well plate were retrieved from the incubator, the culture media were aspirated, and the cells were washed twice with pre-cooled phosphate-buffered saline (PBS). Frozen heart tissue sections were thawed and then immersed in PBS three times, each for 5 minutes. A volume of 1 mL of senescence-associated (SA)-β-gal staining fixative was added and left at room temperature for 15 minutes. The fixative was aspirated, and each well was washed three times with 1 mL of pre-chilled PBS, each for 3 minutes. The PBS was removed, and the working solution was prepared according to the volume of 1 mL of staining solution per well. The ratio of SA-β-galactosidase staining solution A：B：C：X-Gal solution was 1：1：93：5. After wrapping the six-well plate with cling film, it was incubated overnight in a 37°C oven. The following day, photographs were taken under a conventional optical microscope. For cell cultured in plates, the positive rate of SA-β-gal was counted using ImageJ software( http://imagej.nih.gov/ij/); for froze sections, the area of SA-β-gal positive was calculated using ImageJ software.

**Immunofluorescence staining**

Adherent cells and frozen sections of cardiac tissue were stained using a DNA damage detection kit (#C2035S; Beyotime, Nanjing, China). The culture medium of adherent cells cultured in confocal dishes was aspirated and washed twice with PBS. Tissue frozen sections were completely immersed in PBS once. A total of 600 μL of fixative was added and the sections were fixed at room temperature for 15 minutes. The fixative was aspirated and washed with PBS six times, each for 1 minute. A total of 200 μL of blocking solution was added to each dish, and the dishes were left at room temperature for 20 minutes. The blocking solution was aspirated, and 200 μL of ready-use γH2AX, iFluor™ 647 phalloidin （#40762ES75, Yeasen) or p-STAT3(Tyr705) antibody (1:200; #9145, CST, MA, USA) rabbit monoclonal antibody was added to each dish. The dishes were incubated overnight at 4°C. After this incubation, the dishes were washed with PBS six times, each for 3 minutes. A secondary antibody conjugated with fluorescence was added and incubated at room temperature for 1 hour. The dishes were then washed with PBS six times, each for 3 minutes. Mounting medium containing nuclear stain was added, images were captured under a confocal microscope (FV3000; Olympus, Japan).

**Immunoblotting and CO-IP assays**

Total protein from cells or heart tissue was extracted with RIAP buffer (Beyotime). For CO-IP assays, cells were lysed by IP Lysis Buffer and incubated antibodies overnight at 4 °C. The CO-IP kit was used following the manufacturer`s instructions (#C2035S; Beyotime). Nuclear protein was prepared according to the manufacturer`s instructions using NE-PER nuclear cytoplasmic extraction reagent kit (#78833, Thermo Fisher Scientific, Massachusetts, USA). The protein concentration was quantified by the bicinchoninic acid method, and 20–40 μg of protein was subjected to sodium dodecyl sulfate-polyacrylamide gel electrophoresis. Proteins were transferred to a polyvinylidene fluoride (PVDF) membrane, which was washed three times on a Tris-buffered saline with Tween 20 (TBST) shaker for 3–5 minutes each time. Subsequently, the membrane was blocked at room temperature for 1 hour using QuickBlock™ Western Blocking Solution (#P0252; Beyotime). After washing twice with TBST for 3 minutes each time, the PVDF membrane was placed in a preservation box containing the corresponding primary antibodies and incubated overnight at 4°C. Antibody information is as follows: Anti-STAT3 antibody(1:2000, #4904, CST, MA, USA); Anti-Phospho-STAT3(Tyr705) antibody (1:2000; #9145, CST); Anti-Rnd3 antibody (1:1000; #66535-1-Ig, Proteintech, Wuhan, China); Anti-MCP1 antibody(1:1000; #ab214819, Abcam, Cambridge, United Kingdom); Anti-IL-6 antibody (1:1000; #ab259341, Abcam); Anti-IL-1α antibody (1:2000; #DF6893, Affinity, Wuhan, China); Anti-GDF15 antibody (1:2000; #ab206414, Abcam); Anti-p53 antibody(1:2000; #PTM-6319; PTM BIO); Anti-p16(1:2000; #ab51243;Abcam); Anti-β-actin antibody(1:1000; #PTM-5706, PTM BIO), anti-Histone 3(#PTM-6613， PTM BIO). The next day, the PVDF membrane was removed with tweezers and washed three time on a TBST shaker for 3 minutes each time. The secondary antibody was then incubated at room temperature for 1 hour. The PVDF membrane was washed five times with TBST for 3 minutes each time. Enhanced chemiluminescence reagents were added, and the image was developed and photographed with an automatic chemiluminescence imaging system (Tanon-ABLX5 Shanghai, China). The grayscale value was measured using ImageJ.

**Quantitative real-time polymerase chain reaction**

Total RNA was extracted from cells, heart tissue, and serum using the Eastep® Super Reagent Kit (#LS1040; Promega, USA). The Hifair® III 1st Strand cDNA Synthesis SuperMix for quantitative reverse transcription-polymerase chain reaction (RT-PCR) (gDNA digester plus) (#11141ES60; Yeasen, Shanghai, China) was used for the reverse transcription of cDNA, while the Hifair® miRNA 1st Strand cDNA Synthesis Kit (Add A method) (#11148ES10; Yeasen) was used for the reverse transcription of miRNA. The Hieff® qPCR SYBR Green Master Mix (Low Rox Plus) (#11202ES08; Yeasen) was used for RT-PCR. Upon completion of the quantitative RT-PCR run, the melting curves and technical replicates were checked for Ct value discrepancies, ensuring a Ct value difference of <1 and no non-specific amplification before data processing via the Livak method. PCR primers were designed and synthesized by Sangon Biotech (Shanghai, China) and listed below,

| Gene | Specie | Forward primer，5'-3' | Reversed primer，5'-3' |
|---|---|---|---|
| *β-actin* | Rat | CCTGTATGCCTCTGGTCGT | CTGTAGCCACGCTCGGT |
| *Rnd3* | Rat | GCAAGAGCAAACGGAAAG | CATGCCGAAACTAAGGACA |
| *miR-103a-3p* | Rat | GGAGCAGCATTGTACAGGG | CACACTCTCACTCACGCATC |
| *IL-6* | Rat | ACTTCCAGCCAGTTGCCTTCTTG | TGGTCTGTTGTGGGTGGTATCCTC |
| *MCP1* | Rat | CCCTGCTGCCCTTTCTA | CCAATCTGGGGTCACACT |
| *IL-1α* | Rat | CCACATCCCTGTTACCTGA | TGACACCCTGGTTTGAGAA |
| *P53* | Rat | GCGTTGCTCTGATGGTG | CCGAAAAGTCTGCCTGTC |
| *P16* | Rat | CGAACTCGAGGAGAGCGAT | GCCCATCATCATCACCTGTAT |
| *GDF15* | Rat | GCTGCTTCTGCTGTCATGG | CTGGGTCAGGGGTTGGTT |
| *Rnd3* | Human | AATAGAGTTGAGCCTGTGGG | CTAATGTACTAACATCTGTCCGC |
| *miR-103a-3p* | Human | GGAGCAGCATTGTACAGGG | TTGGGAGGTAGGAGGTTGAT |
| *β-actin* | Human | TCTCCCAAGTCCACACAGG | GGCACGAAGGCTCATCA |

**MicroRNA sequencing**

MicroRNA sequencing was performed using the BGI platform (Shenzhen, China)[2]. AC16 cells were cultured in NG and HG medium for 48 hours. Total RNA was extracted using the MiniBEST Universal RNA Extraction Kit (#9767; Takara, Japan), followed by the electrophoresis of RNA samples on polyacrylamide gels, and the recovery of 18–30 nt small RNA (sRNA) from the gels. The corresponding reaction system was set up, and the program was set to link the 3' and 5' ends, reverse transcription, and PCR amplification. The amplified product was subjected to polyacrylamide gel electrophoresis again, and the target band of the library was recovered from the gel and preserved in Ethidium bromide buffer.

The library was quality checked using the appropriate method according to product requirements. The PCR product was denatured into a single strand, a circularization procedure was performed to construct a single-strand circular product, and linear DNA molecules were digested. Rolling circle replication of single-strand circular DNA was performed to prepare DNA nanoball. The DNA nanoball was dripped into the high-density DNA nanochip hole, and sequencing was performed using combined probe anchoring polymerization technology. The raw data obtained from sequencing were filtered. Bowtie2 (v2.2.5) was used to align the clean data to the location on the reference genome to predict miRNA. Subsequently, reads were aligned with miRbase, Rfam database (through cmsearch software), and other databases to annotate the sequencing data for non-coding RNA. To annotate each sRNA with only one annotation, sRNAs were annotated one by one in the following order: miRbase > pirnabank > snoRNA > Rfam > other sRNAs. The Dr. Tom system predicted novel miRNA through miRDeep2 software. Differential gene detection was performed using DEGseq (Q-value ≤0.05 or false discovery rate ≤0.001). RNAhybrid, miRanda, and TargetScan software were used to predict miRNA target genes. The target genes predicted by all three databases were selected, and Phyper was used to perform gene ontology and Kyoto Encyclopedia of Genes and Genomes enrichment analysis on the target genes of differential miRNA. The Bonferroni method was used to correct the candidate genes that met a corrected P value ≤0.05, which was considered significantly enriched.

**Dual luciferase experiment**

Firefly/Renilla luciferase assay [3] was adopted to determine Rnd3 3’UTR promoter activity. 293T cells plated in 24-well plates were co-transfected with 0.1μg Rnd3 3’UTR, 0.4 μg miR-103a-3p-1 (miR-103a-3p precursor) and 0.4 μg negative control (NC) using X-tremegene HP regents (Cat:06366236001, Roche, Switzerland). At the same time, cells transfected with 0.4 μg hsa-mir-146b and 0.1 μg TRAF6 3’UTR were set as positive control. The cells were incubated with DMEM medium plus 10% FBS. 48 hours later post transfection, the luciferase activity (firefly/renilla luminescence ratio) was measured as per the instructions of the dual luciferase reporter system (Cat: #E1910, Promega, USA). All plasmids were constructed by Shanghai Genechem Company.

**Statistical analysis**

GraphPad Prism 9 (GraphPad Software, La Jolla, CA, USA) was used for statistical analysis and graphing of the data. The Shapiro–Wilk test and the QQ plot were used for analysis of the distribution of data. Data with a normal distribution are shown as the mean ± standard deviation (SD, and the unpaired t-test or one-way analysis of variance was used to determine the differences between two or more groups. The Bonferroni post-hoc test was performed when necessary. Data that did not follow a normal distribution are shown as the median ± interquartile range, and the non-parametric Mann–Whitney test was used for comparison between groups. Spearman's and Pearson correlation analysis were used for clinical paraments of clinical cohorts. $P<0.05$ was considered statistically significant.


**Supplemental Figures and Figure Legends**

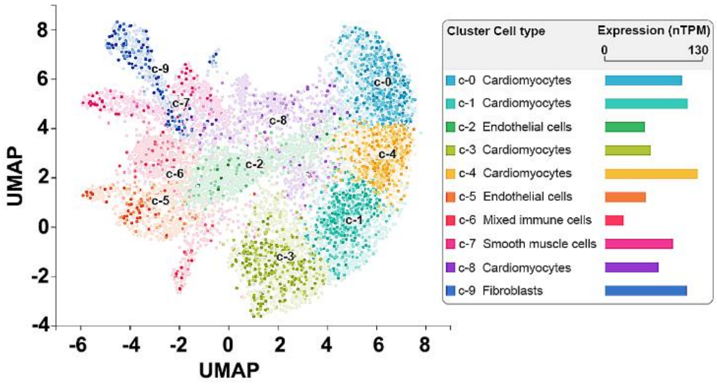


**Figure S1. Expression of Rnd3 mRNA in normal human heart tissue.** Single-cell RNA sequencing shows Rnd3 mRNA levels in normal human cardiac cell types based on the Human Protein Atlas database.

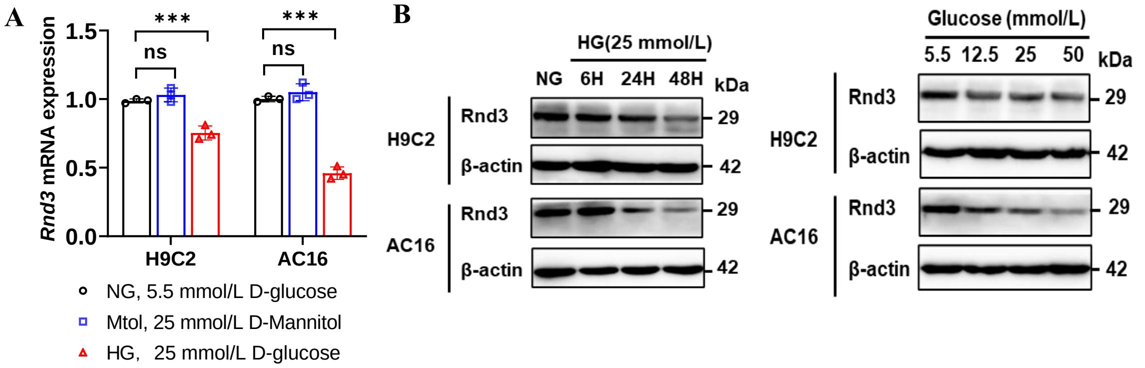


**Figure S2. Detection of *Rnd3* expression in cardiomyocytes**. (**A**) H9C2 and AC16 cells were treated by NG, Mtol and HG condition for 24 hours, then cells were harvested for RT-qPCR analysis of *Rnd3* mRNA. ns, no significance, ****P*<0.001. (**B**) H9C2 and AC16 cells were treated by NG and HG condition for different periods or concentrations, then cells were harvested for western blotting analysis of Rnd3 Protein.

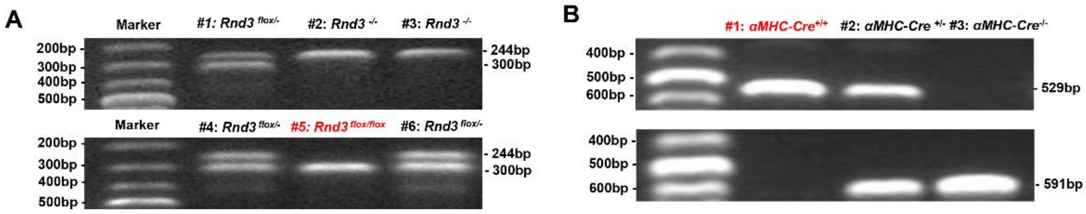


**Figure S3. Genotyping of model rats.** (**A**) PCR was performed using the following primers to identify the genotype of Flox rats. Among them, #5 (red) represents $Flox^{+/+}$ rats. Forward primer (F): 5'-GCA CCT ATG TAG AAG TCC AGG CTT G-3', Reverse primer (R): 5'-AAC TAA GAA GGA CCC TTT GAT CTA CC-3'. (**B**) PCR was performed using the following primers to identify the genotype of cardiomyocyte specific Cre expressing rats. Among them, #1 (red) represents $Cre^{+/+}$ rats. Forward primer(F): 5'- ATT CCT CCT TGA GTT GTG GCA CT-3', Reverse primer_1 (R1): 5'- TGG GCA TGT CTT CAA TCT ACC TC-3'. Reverse primer_2 (R2): 5'-ATG AAC AAA GGT TGG CTA TAA AGA G-3'. Finally, the cardiomyocyte specific *Rnd3* gene knockout rats were obtained by co-breeding # 5 rat in (A) and # 1 rat in (B), and male offspring rats were used for related experiments.

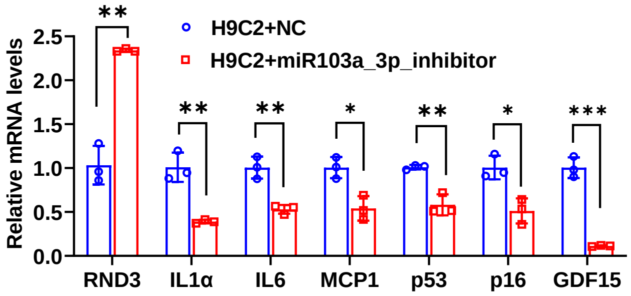


**Figure S4. RT-qPCR analysis of mRNA levels of cellular senescence markers in miR-103a-3p inhibitor treated H9C2 cells**. Unpaired t test, **P*<0.05, ***P*<0.01, ****P*<0.001.

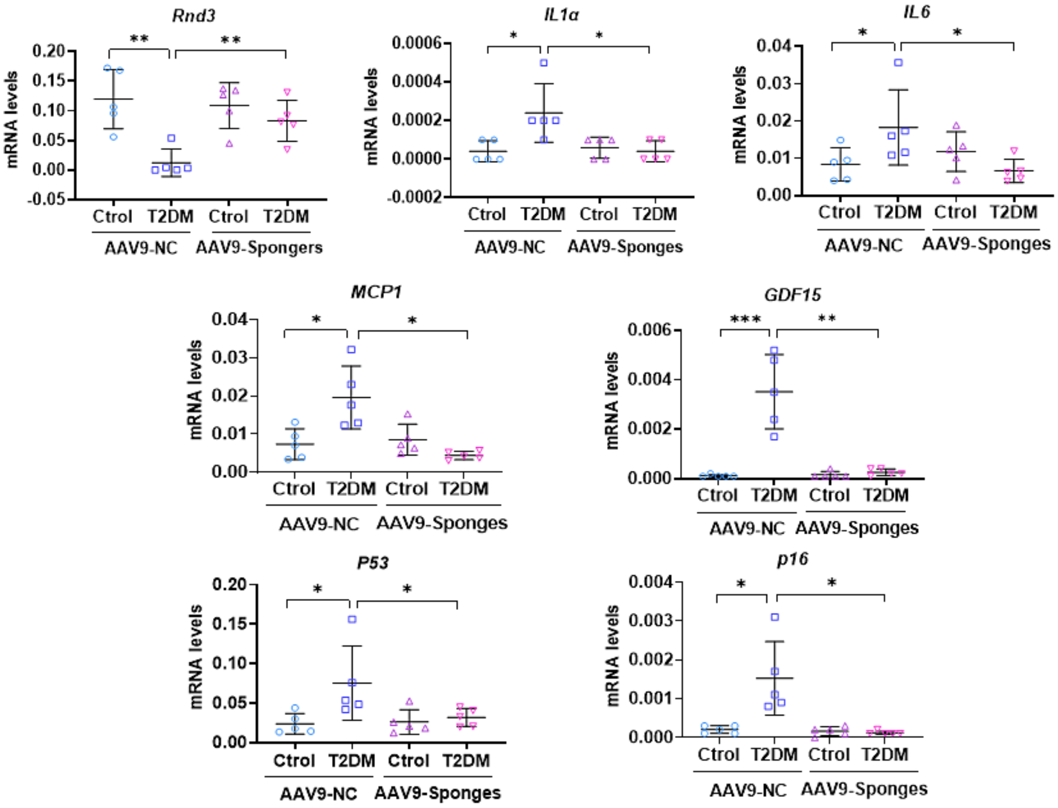


**Figure S5. RT-qPCR was used to analyze the expression of mRNA of *Rnd3* and cellular senescence markers**. Diabetic and non-diabetic rats were injected with AAV9-miR-103a-3p sponges through the tail vein, and the control group was injected with the same titer of AAV9-NC virus. After 8 weeks, cardiac tissues were used qPCR analysis of the expression of mRNA of *Rnd3* and cellular senescence markers. Data were processed by Livak method and expressed as mean ± standard deviation (*n* =5). **P*<0.05, ***P*<0.01, ****P*<0.001.

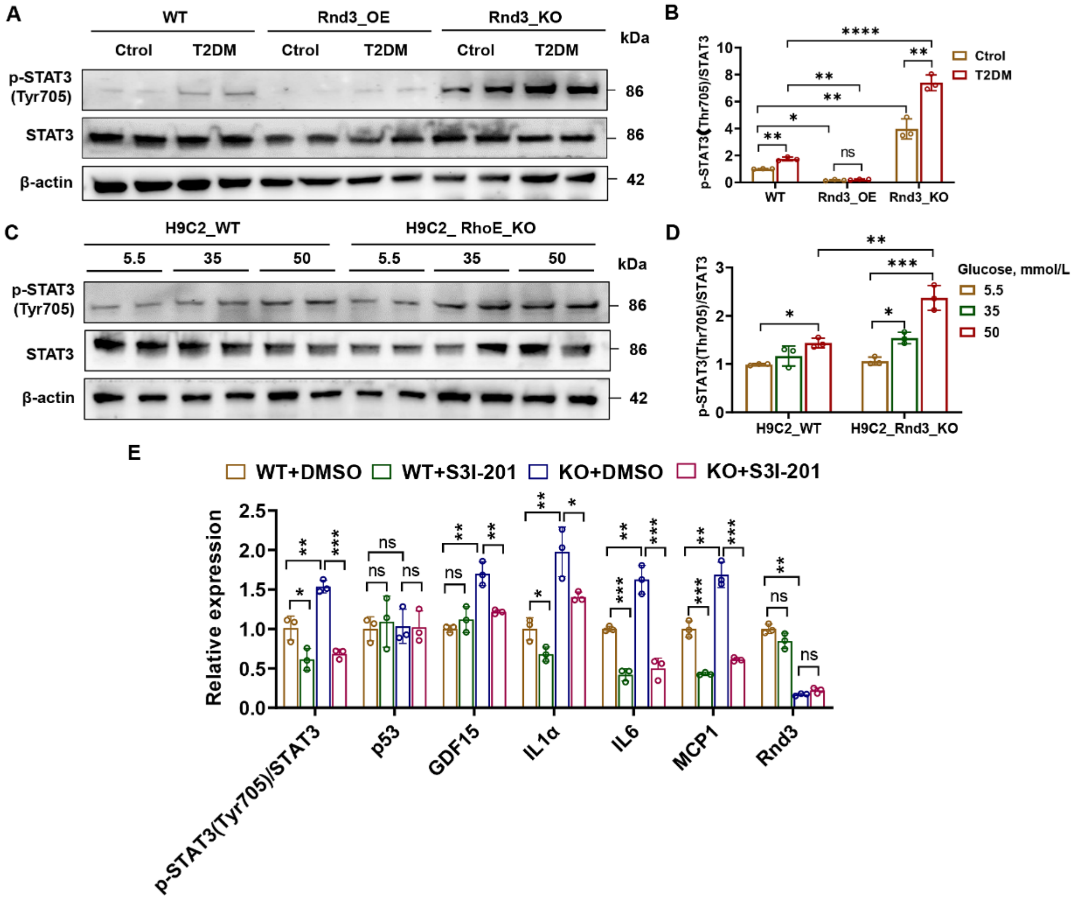


**Figure S6. Effects of diabetes mellitus or HG on STAT3 phosphorylation in cardiac tissues and cardiomyocytes.** (**A**, **B**) Western blot detection of p-STAT3 (Tyr705) and STAT3 protein levels in heart tissue of rats with or without *Rnd3* gene intervention. β-actin served as an internal reference. Data were analyzed by the unpaired t test; *$P$<0.05, **$P$<0.01, ****$P$<0.0001. ns, no significance. (**C**, **D**) Western blot detection of p-STAT3 (Tyr705) and STAT3 protein levels in HG-treated H9C2 cells. β-actin served as an internal reference. Data were analyzed by the unpaired t test; *$P$<0.05, **$P$<0.01, ***$P$<0.001. (**E**) Quantitative analysis of western blot detection of the STAT3 inhibitor S3I-201 on STAT3 activation and cellular senescence in HG-treated H9C2 cells. Data were analyzed by the unpaired t test; *$P$<0.05, **$P$<0.01, ***$P$<0.001, ns, no significance.

**Table S1. Characteristics of clinical cohorts involved in *Rnd3* mRNA test**

| Variate | Overview (n=33) | Control (n=15) | Diabetes (n=18) | p-Value |
|---|---|---|---|---|
| Female, n(%) | 11(33.333) | 6(40.000) | 5(27.778) | 0.458 |
| Man, n(%) | 22(66.667) | 9(60.000) | 13(72.222) | |
| *Rnd3* mRNA, median[IQR] | 0.001[0.000,0.008] | 0.004[0.001,0.009] | 0.000[0.000,0.003] | 0.019 |
| Age, median[IQR] | 59.788±10.525 | 56.867±10.506 | 62.222±9.903 | 0.155 |
| FBG(mmol/l), median[IQR] | 6.020[4.940,9.620] | 4.680[4.180,5.290] | 8.240[7.610,11.870] | <0.001 |
| BUN(mmol/l), median[IQR] | 5.960[4.550,7.600] | 5.060[4.600,6.710] | 6.900[5.960,7.300] | 0.330 |
| CREA(μmol/l), median[IQR] | 73.900[61.600,110.200] | 70.400[65.500,102.600] | 78.800[62.700,110.200] | 0.868 |
| BUN/CR, median[IQR] | 0.070[0.060,0.090] | 0.070[0.060,0.090] | 0.070[0.060,0.090] | 1.000 |
| CK(U/l), median[IQR] | 118.000[84.000,238.000] | 103.000[80.000,181.000] | 169.000[107.000,238.000] | 0.384 |
| CKMB(U/l), median[IQR] | 13.500[7.000,46.200] | 11.300[7.000,37.500] | 14.500[9.330,61.400] | 0.239 |
| MYO（μg/l) ,median[IQR] | 43.160[30.000,75.290] | 30.000[30.000,55.260] | 59.600[33.230,120.970] | 0.023 |
| CTnI(ng/l), median[IQR] | 0.018[0.010,6.330] | 0.010[0.006,0.290] | 0.060[0.010,6.330] | 0.181 |
| NT-BNP(g/l), median[IQR] | 756.000[119.000,1236.000] | 215.000[50.000,756.000] | 1182.000[383.000,2420.000] | 0.014 |
| AST(U/l), median[IQR] | 28.000[23.000,73.000] | 24.000[22.000,36.000] | 44.000[26.000,147.000] | 0.089 |
| LDH(mmol/l), median[IQR] | 228.000[195.000,643.000] | 213.000[195.000,278.000] | 253.000[195.000,665.000] | 0.233 |
| ALT(U/l）, median[IQR] | 25.000[21.000,40.000] | 23.000[21.000,25.000] | 33.000[24.000,55.830] | 0.024 |
| Lpa(mg/l) ,median[IQR] | 155.420[98.830,313.200] | 156.000[123.380,313.200] | 113.800[75.290,298.340] | 0.262 |
| CHOL(mmol/l), mean(±SD) | 4.460[3.550,4.960] | 4.470[4.200,4.820] | 4.160[3.480,4.970] | 0.426 |
| TG(mmol/l), median[IQR] | 1.400[1.020,2.330] | 1.310[1.020,1.770] | 1.400[1.190,2.330] | 0.828 |
| HDL(mmol/l), median[IQR] | 1.116±0.328 | 1.161±0.365 | 1.078±0.287 | 0.488 |
| LDL(mmol/l), mean(±SD) | 2.628±0.757 | 2.663±0.718 | 2.599±0.787 | 0.818 |
| APOA(g/l), median[IQR] | 1.283±0.385 | 1.332±0.357 | 1.242±0.401 | 0.519 |
| APOB(g/l), mean(±SD) | 0.980[0.820,1.120] | 0.970[0.870,1.120] | 0.980[0.800,1.120] | 0.575 |
| APOA/APOB, median[IQR] | 1.376±0.511 | 1.426±0.494 | 1.335±0.520 | 0.624 |
| NE#, median[IQR] | 4.857±2.231 | 3.969±1.040 | 5.597±2.650 | 0.029 |
| NE%, mean(±SD) | 61.455±10.079 | 59.573±8.882 | 63.022±10.729 | 0.343 |
| LYM#, median[IQR] | 2.048±0.723 | 1.970±0.741 | 2.113±0.701 | 0.585 |
| LYM%, mean(±SD) | 29.039±10.575 | 29.613±8.457 | 28.561±12.038 | 0.784 |
| MON#, median[IQR] | 0.500[0.340,0.640] | 0.370[0.300,0.600] | 0.530[0.360,0.650] | 0.255 |
| MON%, mean(±SD) | 6.800[5.400,7.700] | 6.800[5.700,7.300] | 6.700[4.800,8.050] | 0.899 |
| EO#, median[IQR] | 0.160[0.050,0.270] | 0.160[0.070,0.210] | 0.170[0.040,0.290] | 1.000 |
| EO%, median[IQR] | 2.200[0.900,3.400] | 2.500[1.100,3.400] | 1.900[0.600,3.200] | 0.587 |
| AO(mm), mean(±SD) | 28.938±3.657 | 28.867±3.981 | 29.000±3.343 | 0.921 |
| LA(mm), median[IQR] | 35.375±6.353 | 33.200±6.058 | 37.294±5.978 | 0.073 |
| RA(mm), mean(±SD) | 32.000[31.000,35.000] | 32.000[32.000,35.000] | 32.000[31.000,35.000] | 0.863 |
| LVD(mm), median[IQR] | 48.375±5.600 | 47.200±4.679 | 49.412±6.117 | 0.280 |
| LVS(mm), median[IQR] | 33.125±5.407 | 30.733±4.919 | 35.235±4.917 | 0.018 |
| EF%, median[IQR] | 58.939±11.165 | 64.467±6.520 | 54.333±12.101 | 0.008 |
| MVE ,mean(±SD) | 0.785±0.330 | 0.882±0.348 | 0.720±0.301 | 0.247 |
| MVA ,mean(±SD) | 0.927±0.238 | 1.002±0.174 | 0.873±0.262 | 0.207 |
| EA ,median[IQR] | 0.740[0.596,1.080] | 0.794[0.627,1.213] | 0.687[0.580,0.830] | 0.429 |
| FS% ,mean(±SD) | 31.364±7.277 | 34.000±5.138 | 29.167±8.029 | 0.133 |

Continuous variables are described by mean and SD or median and first–third quartile (interquartile range, IQR), categorical ones by absolute and relative frequencies. FBG, Fasting blood glucose; BUN, Blood Urea Nitrogen; CREA, Creatinine; CK, Creatine kinase; CK-MB, Cardiac isoenzyme of creatine kinase; LDH, Lactate dehydrogenase; ALT, Alanine aminotransferase; CHOL, Cholesterol; TG, Triglyceride; HDL, High density lipoprotein; LDL, Low Density Lipoprotein; APOA, Apolipoprotein A; APOB, Apolipoprotein B; NE, Neutrophil; LYM, Absolute lymphocyte count; MON Monocyte; EO, Eosinophil; FDP, Fibrinogen degradation products; AO, Aorta diameter; LA, Left atrial diameter; RA, Right atrial diameter; LVD, left ventricular diameter; LVS, Left ventricular end-systolic diameter; EF%, Ejection fraction; AST, Aspartate aminotransferase; cTnI, Cardiac troponin I; NT-proBNP, N-terminal brain natriuretic peptide precursor

**Table S2. Characteristics of T2DM and control rats with or without *Rnd3* gene intervening**

| Variate | Overview (n=30) | WT_Control (n=5) | WT_D2TM (n=5) | KO_Control (n=5) | KO_T2DM (n=5) | OE_Control (n=5) | OE_D2TM (n=5) | p-Value |
|---|---|---|---|---|---|---|---|---|
| 6w FBG(mmol/L), mean(SD) | 8.713 (2.069) | 10.400 (2.304) | 8.200 (1.994) | 8.060 (1.306) | 7.840 (1.970) | 8.860 (2.392) | 8.920 (0.852) | 0.449 |
| 6w Weight(g), mean(SD) | 149.767 (15.830) | 153.200 (10.127) | 164.400 (12.225) | 161.400 (4.499) | 146.800 (13.014) | 134.200 (5.269) | 138.600 (17.235) | 0.004 |
| 10w FBG(mmol/L), mean(SD) | 17.213 (8.910) | 8.940 (1.560) | 26.400 (3.603) | 8.740 (1.401) | 27.240 (0.967) | 8.380 (2.393) | 23.580 (2.926) | <0.001 |
| 10w Weight(g), mean(SD) | 306.333 (44.257) | 294.000 (16.625) | 314.000 (9.695) | 310.600 (18.959) | 342.800 (16.092) | 250.000 (41.593) | 326.600 (62.372) | 0.012 |
| 18w FBG(mmol/L), median[IQR] | 15.210[8.600,31.700] | 8.600[8.000, 8.900] | 33.100[31.700,33.300] | 9.100[7.100, 10.900] | 31.500[29.100,33.000] | 6.700[5.900, 14.500] | 26.400[21.700, 31.900] | <0.001 |
| 18w Weight(g), mean(SD) | 354.960 (42.319) | 379.100 (41.149) | 327.520 (14.580) | 369.800 (11.285) | 392.800 (9.432) | 361.380 (40.157) | 299.160 (29.461) | <0.001 |
| 18w Length of tibia(cm), mean(SD) | 5.267 (0.888) | 5.760 (0.224) | 3.480 (0.256) | 5.620 (0.117) | 5.920 (0.117) | 5.580 (0.458) | 5.240 (0.539) | <0.001 |
| 18w Heart weight(g), mean(SD) | 1.550 (0.269) | 1.480 (0.293) | 1.840 (0.196) | 1.360 (0.206) | 1.740 (0.102) | 1.420 (0.232) | 1.460 (0.136) | 0.014 |

**Table S3. Differential expression of miRNAs enriched by miRNA sequencing**

| Gene ID | log2(HG/NG) | P_value (HG/NG) | Q_value (HG/NG) |
|---|---|---|---|
| hsa-miR-101-5p | 1.45 | 5.13E-03 | 2.84E-02 |
| hsa-miR-103a-3p | 5.51 | 0.00E+00 | 0.00E+00 |
| hsa-miR-103b | -1.59 | 0.00E+00 | 0.00E+00 |
| hsa-miR-107 | 1.05 | 1.94E-10 | 2.22E-09 |
| hsa-miR-1273h-5p | -2.73 | 2.56E-03 | 1.54E-02 |
| hsa-miR-1973 | -1.87 | 2.11E-21 | 3.59E-20 |
| hsa-miR-199b-3p | -5.85 | 0.00E+00 | 0.00E+00 |
| hsa-miR-200a-3p | 1.89 | 4.53E-04 | 3.25E-03 |
| hsa-miR-3163 | -3.29 | 3.54E-03 | 2.03E-02 |
| hsa-miR-3194-3p | 1.45 | 5.63E-07 | 5.40E-06 |
| hsa-miR-33b-5p | -1.38 | 7.88E-07 | 7.51E-06 |
| hsa-miR-3529-3p | 8.49 | 0.00E+00 | 0.00E+00 |
| hsa-miR-365a-3p | -4.75 | 5.70E-64 | 2.16E-62 |
| hsa-miR-3691-5p | 1.90 | 2.12E-03 | 1.30E-02 |
| hsa-miR-3913-5p | 1.72 | 8.69E-03 | 4.40E-02 |
| hsa-miR-423-5p | -1.05 | 0.00E+00 | 0.00E+00 |
| hsa-miR-4463 | -1.61 | 2.41E-05 | 1.97E-04 |
| hsa-miR-4484 | 1.05 | 2.09E-04 | 1.54E-03 |
| hsa-miR-4485-5p | -2.24 | 1.01E-03 | 6.73E-03 |
| hsa-miR-4689 | -3.47 | 1.09E-05 | 9.23E-05 |
| hsa-miR-4732-3p | 3.24 | 1.30E-21 | 2.23E-20 |
| hsa-miR-4804-5p | -1.74 | 1.47E-03 | 9.42E-03 |
| hsa-miR-5090 | 2.24 | 6.10E-03 | 3.28E-02 |
| hsa-miR-548ad-5p | -5.12 | 3.32E-34 | 7.72E-33 |
| hsa-miR-548ae-5p | -1.75 | 9.00E-08 | 8.89E-07 |
| hsa-miR-548ap-3p | 2.20 | 1.40E-22 | 2.48E-21 |
| hsa-miR-548o-5p | -7.43 | 7.64E-04 | 5.23E-03 |
| hsa-miR-6511a-3p | 1.23 | 1.17E-04 | 9.06E-04 |
| hsa-miR-6820-3p | 1.31 | 2.36E-03 | 1.43E-02 |
| hsa-miR-7-5p | -8.57 | 0.00E+00 | 0.00E+00 |
| hsa-miR-7974 | 1.15 | 3.79E-15 | 5.03E-14 |
| hsa-miR-937-5p | 1.20 | 8.95E-04 | 6.03E-03 |
| novel-hsa-miR16-3p | 1.21 | 7.08E-03 | 3.69E-02 |
| novel-hsa-miR184-3p | -2.37 | 4.95E-06 | 4.48E-05 |
| novel-hsa-miR187-5p | 3.55 | 1.16E-03 | 7.61E-03 |
| novel-hsa-miR193-5p | -1.40 | 1.31E-04 | 1.00E-03 |
| novel-hsa-miR243-5p | 1.44 | 1.02E-05 | 8.63E-05 |
| novel-hsa-miR263-3p | -3.29 | 3.54E-03 | 2.03E-02 |
| novel-hsa-miR295-3p | 1.63 | 5.47E-04 | 3.86E-03 |
| novel-hsa-miR300-3p | 1.16 | 7.07E-06 | 6.27E-05 |
| novel-hsa-miR35-5p | -1.04 | 1.36E-04 | 1.04E-03 |
| novel-hsa-miR56-5p | 1.20 | 8.43E-03 | 4.28E-02 |
| novel-hsa-miR99-5p | 2.56 | 5.47E-03 | 2.99E-02 |

**Table S4. Characteristics of clinical cohorts for circulating miR-103a-3p test**

| Variate | Missing | Overview (n=41) | Control (n=17) | Diabetes (n=24) | p-Value |
|---|---|---|---|---|---|
| Female, n(%) | 0 | 14(34.146) | 7(41.176) | 7(29.167) | 0.424 |
| Man, n(%) | | 27(65.854) | 10(58.824) | 17(70.833) | |
| Age ,mean(±SD) | 0 | 62.195±8.244 | 63.118±9.480 | 61.542±7.170 | 0.558 |
| miR-103a-3p, median [IQR] | 0 | 0.258[0.085,0.575] | 0.041[0.016,0.118] | 0.542[0.308,0.612] | <0.001 |
| FBG(mmol/l), median[IQR] | 0 | 5.910[4.760,8.090] | 4.920[4.300,5.810] | 7.140[5.650,9.380] | <0.001 |
| Tyg, median[IQR] | 0 | 1.106[0.863,1.286] | 0.855[0.767,0.969] | 1.278[1.106,1.374] | <0.001 |
| NE#, median[IQR] | 2 | 4.560[3.520,6.000] | 4.780[3.610,6.290] | 4.520[3.400,5.350] | 0.388 |
| NE%, median[IQR] | 2 | 62.700[57.300,70.800] | 68.000[58.100,74.600] | 60.500[54.800,64.200] | 0.183 |
| LYM#, mean(±SD) | 2 | 1.910±0.766 | 1.716±0.739 | 2.060±0.752 | 0.173 |
| LYM%, mean(±SD) | 2 | 26.728±10.086 | 23.788±10.657 | 29.000±8.985 | 0.115 |
| MON#, mean(±SD) | 2 | 0.422±0.133 | 0.419±0.140 | 0.423±0.128 | 0.932 |
| MON%, median[IQR] | 2 | 5.900[4.900,7.000] | 5.100[4.700,7.300] | 5.900[4.900,7.000] | 0.821 |
| EO#, median[IQR] | 2 | 0.110[0.050,0.180] | 0.150[0.050,0.190] | 0.090[0.040,0.150] | 0.487 |
| EO%, median[IQR] | 2 | 1.400[0.600,3.100] | 2.200[0.700,3.100] | 1.200[0.600,2.900] | 0.307 |
| ALT(U/l), median[IQR] | 4 | 21.000[15.000,32.000] | 18.000[14.000,30.000] | 23.000[20.000,34.000] | 0.136 |
| AST(U/l), median[IQR] | 1 | 21.000[17.000,24.000] | 21.000[17.000,24.000] | 21.000[17.000,24.000] | 0.670 |
| ALP(U/l), median[IQR] | 16 | 73.000[63.000,86.000] | 73.000[68.000,82.000] | 74.000[59.000,86.000] | 1.000 |
| ALB(g/l), median[IQR] | 4 | 36.200[34.600,38.900] | 35.100[33.400,37.000] | 37.200[36.000,39.600] | 0.061 |
| TC(mmol/l), median[IQR] | 0 | 4.650[3.750,5.380] | 3.930[3.620,4.650] | 5.220[4.180,5.830] | 0.039 |
| TG(mmol/l), median[IQR] | 0 | 1.410[1.050,1.930] | 1.190[0.980,1.530] | 1.820[1.350,2.100] | 0.020 |
| HDL(mmol/l),m ean(±SD) | 0 | 1.079±0.244 | 1.147±0.255 | 1.031±0.223 | 0.141 |
| LDL(mmol/l), median[IQR] | 0 | 2.790[1.980,3.670] | 2.220[1.980,2.790] | 3.510[2.390,3.890] | 0.046 |
| Lp(a)(mg/l), mean(±SD) | 0 | 1.136±0.200 | 1.165±0.236 | 1.116±0.167 | 0.453 |
| APOB(g/l), median[IQR] | 0 | 0.990[0.770,1.190] | 0.900[0.730,1.030] | 1.160[0.920,1.320] | 0.078 |
| BUN(μmol/l), mean(±SD) | 3 | 5.403±1.453 | 5.412±1.613 | 5.396±1.326 | 0.974 |
| Cr(μmol/l), median[IQR] | 3 | 70.500[56.800,83.900] | 72.700[61.500,106.000] | 70.500[51.500,82.500] | 0.367 |
| UA(μmol/l）, median[IQR] | 3 | 355.000[285.000,417.000] | 367.000[285.000,375.000] | 355.000[287.000,443.000] | 0.636 |

Continuous variables are described by mean and SD or median and first–third quartile (interquartile range, IQR), categorical ones by absolute and relative frequencies. FBG, Fasting blood glucose; Tyg, Triglyceride-glucose index; ALT, Alanine aminotransferase; AST, Aspartate aminotransferase; ALP, Alkaline phosphatase; ALB, Albumin; TC, Total cholesterol; TG, Triglyceride; HDL, High density lipoprotein; LDL, Low Density Lipoprotein; Lp(a), Lipoprotein a; APOB, Apolipoprotein B; BUN, Blood Urea Nitrogen; Cr, Creatinine; UA, Serum trioxypurine.

**Table S5. Characteristics of T2DM and control rats with or without AAV9_miR103a-3p_sponges infusing**

| Variate | Overview (n=20) | WT_Control (n=5) | WT_T2DM (n=5) | OE_Control (n=5) | OE_T2DM (n=5) | p-Value |
|---|---|---|---|---|---|---|
| 6w FBG(mmol/l), mean(SD) | 8.265 (1.863) | 8.600 (1.231) | 7.240 (1.332) | 8.500 (2.273) | 8.720 (2.000) | 0.618 |
| 6w Weight(g), mean(SD) | 176.500 (12.476) | 170.000 (9.940) | 180.600 (12.110) | 174.600 (12.595) | 180.800 (11.720) | 0.512 |
| 10w FBG (mmol/l), mean(SD) | 18.140 (9.187) | 9.220 (0.571) | 27.380 (3.666) | 9.700 (1.149) | 26.260 (4.514) | <0.001 |
| 10w Weightg(g), mean(SD) | 270.700 (55.484) | 234.800 (43.398) | 246.000 (30.679) | 289.800 (33.855) | 312.200 (66.010) | 0.092 |
| 18w FBG (mmol/l), median[IQR] | 19.500[9.600, 24.500] | 9.400[8.100, 10.700] | 22.600[21.000, 33.300] | 9.600[9.000, 9.700] | 25.900[24.500, 28.100] | 0.002 |
| 18w Weight(g), mean(SD) | 313.235 (66.039) | 296.260 (38.196) | 294.860 (42.249) | 324.460 (41.564) | 337.360 (105.544) | 0.726 |
| 18w Length of tibia(cm), mean(SD) | 4.875 (0.440) | 4.720 (0.412) | 4.980 (0.232) | 4.820 (0.397) | 4.980 (0.588) | 0.782 |
| 18w Heart weight(g), mean(SD) | 1.488 (0.288) | 1.318 (0.173) | 1.778 (0.066) | 1.557 (0.058) | 1.299 (0.375) | 0.017 |
